# Association between bullying, state-level policy and mental health symptoms in gender diverse youth

**DOI:** 10.64898/2025.12.19.25342709

**Authors:** Dylan E. Hughes, Sarah L. Zapetis, Arianna Mordy, Daisy Lopez, Vanessa Calderon, Laura Adery, Rachel Martino, Sarah E. Chang, Lucina Q. Uddin, Carlos Cardenas-Iniguez, Richard T. Lebeau, Natalia Ramos, Lauren C. Ng, Katherine H. Karlsgodt, Carrie E. Bearden

## Abstract

**Importance:** As the percentage of young people in the United States identifying as transgender and gender diverse increases, more youth face identity-linked social and structural stigma and discrimination. Little is known about the impact of stigma on psychotic-like experiences in gender diverse youth.

**Objective:** To assess the impact of bullying victimization and state-level policies on psychotic-like experiences among gender diverse youth.

**Design:** In this prospective cohort study, cross-sectional and longitudinal analyses were conducted using data collected between 2017-2022 across 17 states.

**Setting:** The Adolescent Brain Cognitive Development (ABCD) Study is a U.S. population-based longitudinal study that follows and deeply phenotypes adolescents from the age of 9 to 18.

**Participants:** Cross-sectional analyses included data from 9,112 participants (mean age=13 ± 0.6) collected between 2019 and 2022. Longitudinal analyses comprised 4,529 participants with data collected across 5 waves between 2017 and 2022.

**Exposures:** Self-reported frequency of bullying victimization and data on annual state-level policies related to gender identity.

**Main Outcomes:** Self-reported psychotic-like experiences and associated distress, measured by the Prodromal Questionnaire - Brief Child Version.

**Results:** Based on a dimensional measure of gender, 689 adolescents were identified as most gender diverse (i.e., least congruent with birth-assigned sex) and 8,240 as least gender diverse (i.e., most congruent with birth-assigned sex). Rates of bullying victimization and psychotic-like experiences were significantly elevated in the most vs. least gender diverse group, with bullying partially mediating the difference in psychotic-like experiences (indirect effect = 0.11, p < 2×10^-16^; direct effect = 0.52, p < 1×10^-16^). Gender diverse adolescents exhibited greater sensitivity to the effects of bullying on psychotic-like experiences (interaction ß = 0.14, 95% CI [0.09, 0.19], p = 8.5×10^-08^). Moreover, the persistence of unsupportive legislation across 4 years was associated with significantly greater increases in psychotic-like experiences over time in gender diverse youth (interaction ß = 0.30, 95% CI [0.20, 0.40], p = 2.2×10^-8^).

**Conclusions:** These findings indicate that bullying victimization and unsupportive legislation may explain greater and increasing rates of psychotic-like experiences in gender diverse youth.

**KEY POINTS:** *Question:* Do bullying and state-level policy related to gender identity contribute to mental health problems in gender diverse youth in the United States?

*Finding:* In this large U.S.-based sample of adolescents (ages 9-13), gender diverse adolescents reported more frequent experiences of bullying, which partially accounted for increased rates of subclinical psychotic-like experiences (PLEs). Between 2017 and 2022, gender diversity was associated with increasing PLEs only in states with consistently unsupportive policies; in all other states, PLE scores remained stable over time or decreased.

*Meaning:* Results suggest that PLEs in the context of gender diversity are partially attributable to the sociopolitical environment and that policy decisions at the state-level have far-reaching impacts on the mental health of youth in the United States.

## INTRODUCTION

Between 2017 and 2022, the percentage of youth (ages 13-17) identifying as transgender and gender diverse (TGD) in the United States doubled from 0.73%^1^ to 1.43%^2^. As gender is increasingly reported on a spectrum^3,4^, these numbers may underestimate the proportion of the United States (U.S.) adolescent population whose gender identity is dimensionally incongruent with their birth-assigned sex (e.g., a birth-assigned male who does not feel entirely like a boy). As gender becomes increasingly nuanced, there is a growing need to understand the mechanisms underlying the disproportionate prevalence of mental health problems among TGD youth^5–7^ and whether dimensionally incongruent youth are vulnerable to the same effects.

Psychotic spectrum symptoms in particular are overrepresented in TGD individuals. TGD adults are 3 to 49.7 times more likely to have a schizophrenia spectrum disorder diagnosis than cisgender adults^8^. In TGD youth, estimates for the prevalence of psychotic-like experiences (PLEs) – psychotic spectrum symptoms that may be accompanied by distress and/or indicate risk for clinically significant psychosis^9^ – are sparse. Still, limited evidence suggests that TGD youth (ages 12-28) are overrepresented in samples at high risk for clinically significant psychosis^10^. Nevertheless, the etiology of psychosis across its continuum in TGD youth remains understudied^11^.

The minority stress model^12^ provides a framework to examine the mechanisms through which minoritized groups, including TGD youth, experience higher rates of mental health problems like psychosis. The model proposes that chronic exposure to external (i.e., distal) stressors – such as discrimination and rejection – shapes negative attitudes towards one’s identity (i.e., proximal stressors), which in turn may lead to mental health challenges^11,12^. In line with this framework, TGD youth experience high rates of, and are more vulnerable to, peer victimization, discrimination, and other interpersonal stressors^6,13,14^.

Structural stigma (i.e., public attitudes, societal conditions, and/or policies that explicitly or implicitly impose barriers to the well-being of minoritized groups) is another form of distal stress that can negatively impact mental health in minoritized individuals^15–17^. In TGD youth in the U.S., supportive policies (e.g., anti-discrimination laws in housing, school, and public spaces) have been shown to protect against mental health problems^18,19^. Unsupportive policies (e.g., “bathroom laws” that block transgender people from using public spaces aligned with their gender identity) are associated with poorer mental health^20^. Alarmingly, one U.S. study showed that, following the enactment of unsupportive laws between 2018-2022, the incidence of suicide attempts among TGD youth increased by 7% to 72%^20^. This association is particularly concerning given the proliferation of legislation targeting lesbian, gay, bisexual, transgender, and queer/questioning (LGBTQ+) Americans; as of June 2025, 588 anti-LGBTQ+ bills had been introduced across the U.S. in 2025, twice as many as were introduced throughout the entire year of 2022^21^.

Hypervigilance and paranoid ideation are core features of psychosis^22^ and may be engendered by exposure to the unique interpersonal stressors and structural stigma experienced by minoritized groups^23^. While interpersonal stressors such as bullying have been shown to partially explain high rates of PLEs in youth with diverse expressions of gender^24^, it is unclear whether this is evident in youth with marginally incongruent gender identities. Few, if any, studies, have reported on the effects of policy on PLEs in TGD youth.

In this study, we explore the effects of two distinct forms of distal stressors—interpersonal (i.e., bullying) and structural (i.e., a lack of supportive state-level policies related to gender-identity)—on psychotic-like experiences (PLEs) in a large adolescent dataset collected across 17 states. We hypothesized that (1) bullying would mediate the relationship between gender diversity and PLEs, (2) the effect of bullying on PLEs would be stronger in the most gender-diverse youth compared to non-gender diverse youth, and (3) the absence of supportive legislation would be associated with more PLEs in gender-diverse youth.

## METHODS

### Data

The analytic sample was derived from 11,868 youth enrolled in the Adolescent Brain Cognitive Development (ABCD) Study (Data Release 5.1), a population-based, prospective longitudinal study across 21 U.S. sites (17 states) following adolescents annually from age 9-18. Participants were on average 10.9 ± 0.64 (s.d.) years old (47.9% female) at the first timepoint (Year 1) and 14.1 ± 0.68 years old (47.5% female) at the last available timepoint (Year 4). State-level legislative data were provided by the Movement Advancement Project (MAP)^25^, an organization that collates data related to LGTBQ+ rights in the U.S. Cross-sectional analyses were performed on the Year 3 data (modal interview year = 2020), the largest quantity of data (n = 10,123) at the oldest age (mean age = 12.9). Longitudinal analyses were conducted across Years 1 (n = 11,220; mean age = 10.9; modal year = 2018) through 4 (n = 4,688; mean age = 14.1; modal year = 2021).

### Gender diversity

Gender diversity was operationalized using responses to two questions on a gender questionnaire constructed by ABCD to measure felt-gender^4^: “How much do you feel like a boy?” and “How much do you feel like a girl?” with response options on a 5-point Likert scale ranging from “Not at all” to “Totally”. At each timepoint, cutoff points were applied to the scores to create four groups at each timepoint, ranging from the least gender diverse (i.e., felt-gender most congruent with birth-assigned sex) to most gender diverse (i.e., felt-gender least congruent with birth-assigned sex) (**Figure 1**, **Supplementary Table 1**). Unless otherwise noted, the effects of group 4 (most gender diverse) versus group 1 (least gender diverse; referent) were the primary effects interpreted.

**Figure 1.**
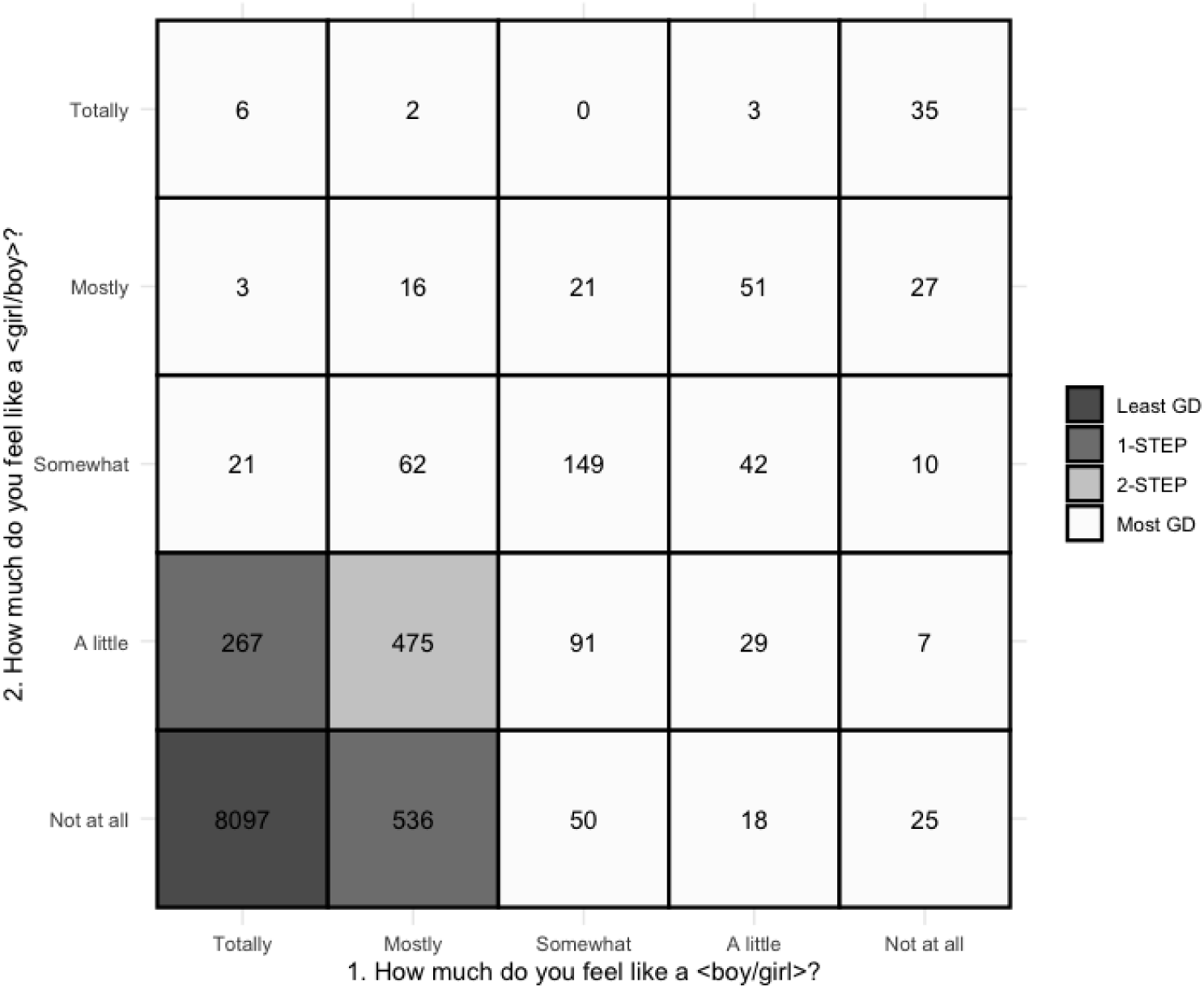
Operationalization of gender diversity variable from the two felt-gender questions at Year 3 data, the timepoint at which cross-sectional analyses were conducted. Participants that answered that they felt ‘totally’ like the gender associated with their birth-assigned sex and ‘not at all’ like the other gender were categorized in the least gender diverse group (shown in dark grey; Least GD; n = 8,240). Participants who responded that they felt ‘somewhat’, ‘a little’, or ‘not at all’ like their birth-assigned gender (question 1, x-axis) and ‘somewhat’, ‘mostly’, or ‘totally’ like the other gender (question 2, y-axis) were categorized in the most gender diverse group (shown in white; Most GD; n = 689). Adapted from Potter et al. 2021.

### Psychotic-like experiences (PLEs)

PLEs were measured via the Prodromal Questionnaire - Brief Child Version (PQ-BC), a 21-item developmentally appropriate self-report measure of psychotic-like experiences, and associated distress, using a 5-point Likert scale. The distress-weighted sum of the responses to all items was used in analyses, as done previously^26,27^ (see **Supplemental Methods**).

### Broad mental health problems

The Brief Problem Monitor (BPM) total T-score was used as a measure of general psychopathology. The BPM is a 19-item self-report measure that assesses mental health problems across multiple domains, where the total score is the scaled sum of all items^28^.

### Bullying victimization and perpetration

Participants reported on the frequency of bullying experiences via a 9-item questionnaire, the Peer Experiences Questionnaire (PEQ), with each item endorsed on a scale of 1 (never) to 5 (a few times a week). The PEQ consists of two versions, one inquiring about bullying victimization and the other perpetration. The sum of all 9 victimization items was used for primary analyses, with greater scores representing greater rates of victimization. The perpetration scale was included as a covariate in sensitivity analyses, as bullying victimization is correlated with perpetration^29^.

### State-level policies related to gender identity or expression

Tallies of enacted protective and anti-transgender legislation for all US states across all years of data collection (2017-2022) were provided by MAP^25^, which assigns each state a score based on the number of enacted laws related to LGTBQ+ rights. In the current analysis, the gender-identity-related MAP tallies were analyzed, representing tallies of laws that “explicitly address or impact gender identity and/or expression”^25^. Tallies were converted to a proportion based on the maximum tally for a given year. Thus, each participant was assigned a value, wherein a score closer to 1 represents a participant residing in a state with more supportive gender identity policies, and a score closer to 0 represents a state with less supportive policies. For longitudinal analyses, as most state scores varied minimally across the 4 years, states were categorized as having consistently high (i.e., scores above 0.5 across time), consistently low (scores below 0.5), or increasingly supportive policies (see **Supplemental Methods** for detail).

### Statistical Analysis

Using R Statistical Software (v4.3.1)^30^, cross-sectional analyses were conducted with linear mixed effects regression (*lme4* package) with families nested within sites. Mediation models were conducted with the *mediation* package in R. Because this package does not support complex nested designs, a random intercept was allowed only for family ID; site was included as a fixed effect. To test the moderating effect of gender diversity on the relationship between bullying victimization and PLEs, an interaction term between bullying victimization and gender diversity was included. Longitudinal analyses allowed the coefficient of the main independent variable to vary by time within subject, within family, and within site (i.e., random slope by time). Additionally, to test whether change in PLE scores over time differed by gender diversity (main effect: most versus least gender diverse) and/or state policy, interaction terms between the variable(s) of interest and time were included. All models were repeated with broad mental health problems as the dependent variable to test whether any observed significant effects were specific to PLEs. All reported cross-sectional models included age, pubertal development, birth-assigned sex, parental education, and combined family income as fixed-effects covariates. Supplementary analyses included race and ethnicity as covariates^31,32^ (see Supplement). Longitudinal models allowed a random slope by time and included the same covariates but omitted age as it was collinear with time. Models examining effects of state-level policies covaried for a state’s Gini coefficient, a time-variant measure of state-level income inequality (see Supplement for details about covariates).

## RESULTS

On average, adolescents (see **Table 1** for demographics) in the most gender diverse group scored 0.63 standard deviations higher on the measure of PLEs (PQ-BC) than their least gender diverse peers (ß = 0.63, 95% CI [0.56, 0.70], p < 1×10^-16^; **Supplementary Tables 2, 3**). Additionally, the most gender diverse group endorsed more broad mental health problems (BPM; ß=0.90, 95% CI [0.80, 0.99], p<1×10^-16^; **Supplementary Table 4**) and bullying victimization (PEQ-Vic; ß=0.48, 95% CI [0.39, 0.56], p<1×10^-16^; **Supplementary Tables 5, 6**); the latter effect remained significant after controlling for bullying perpetration (ß = 0.36, 95% CI [0.28, 0.43], p<1×10^-16^; **Supplementary Table 7**).

**Table 1.** Demographics at Year 3 data collection.

| **Variable** | **Levels** | **Least GD**<br>**(n=8,240)** | **1-STEP**<br>**(n=825)** | **2-STEP**<br>**(n=491)** | **Most GD**<br>**(n=689)** | **P** |
| --- | --- | --- | --- | --- | --- | --- |
| Age | Mean (SD) | 12.9 (0.6) | 12.9 (0.7) | 12.8 (0.6) | 12.9 (0.6) | <0.001 |
| Birth-Assigned Sex | F | 3289 (40.6) | 543 (67.6) | 381 (80.2) | 549 (82.2) | <0.001 |
|  | M | 4808 (59.4) | 260 (32.4) | 94 (19.8) | 119 (17.8) |  |
| Pubertal Development | Mean (SD) | 2.5 (0.7) | 2.7 (0.7) | 2.9 (0.6) | 2.9 (0.6) | <0.001 |
| Race | White | 5487 (67.5) | 517 (63.8) | 311 (63.7) | 394 (58.2) | <0.001 |
|  | Black | 1098 (13.5) | 132 (16.3) | 61 (12.5) | 125 (18.5) |  |
|  | Asian | 198 (2.4) | 23 (2.8) | 9 (1.8) | 4 (0.6) |  |
|  | AIAN/NHPI | 49 (0.6) | 6 (0.7) | 5 (1.0) | 4 (0.6) |  |
|  | Other | 345 (4.2) | 37 (4.6) | 23 (4.7) | 30 (4.4) |  |
|  | Mixed | 953 (11.7) | 95 (11.7) | 79 (16.2) | 120 (17.7) |  |
| Ethnicity | Non-Hispanic | 6513 (80.0) | 645 (79.2) | 393 (81.5) | 510 (75.2) | 0.019 |
|  | Hispanic | 1629 (20.0) | 169 (20.8) | 89 (18.5) | 168 (24.8) |  |
| Parental Education | Mean (SD) | 17.4 (2.5) | 17.1 (2.8) | 17.3 (2.5) | 16.7 (2.6) | <0.001 |
| Family Income | Mean (SD) | 7.6 (2.3) | 7.3 (2.4) | 7.4 (2.2) | 6.8 (2.5) | <0.001 |
| Psychotic-like Experiences | Mean (SD) | 2.1 (5.3) | 4.5 (8.3) | 6.3 (9.8) | 7.7 (11.1) | <0.001 |
| Broad mental health problems | Mean (SD) | 53.4 (5.2) | 56.2 (6.5) | 58.2 (6.9) | 59.1 (7.3) | <0.001 |
| Bullying victimization | Mean (SD) | 2.7 (3.6) | 3.3 (3.9) | 4.4 (4.9) | 4.5 (4.9) | <0.001 |
| Bullying perpetration | Mean (SD) | 1.1 (2.0) | 1.4 (2.2) | 1.6 (2.5) | 1.4 (2.2) | <0.001 |

### Mediating effect of bullying victimization

In the mediation analysis, the indirect effect of gender diversity on PLEs through bullying victimization was significant, with 17% of the total effect mediated by bullying victimization (indirect effect = 0.11, p < 2×10^-16^, direct effect = 0.53, p < 1×10^-16^; **Figure 2**). Likewise, the difference in broad mental health problems by gender diversity was partially mediated by bullying victimization (15% mediated, indirect effect = 0.14, p < 2×10^-16^, direct effect = 0.80, p < 2×10^-16^; **Supplementary Figure 1**).

**Figure 2.**
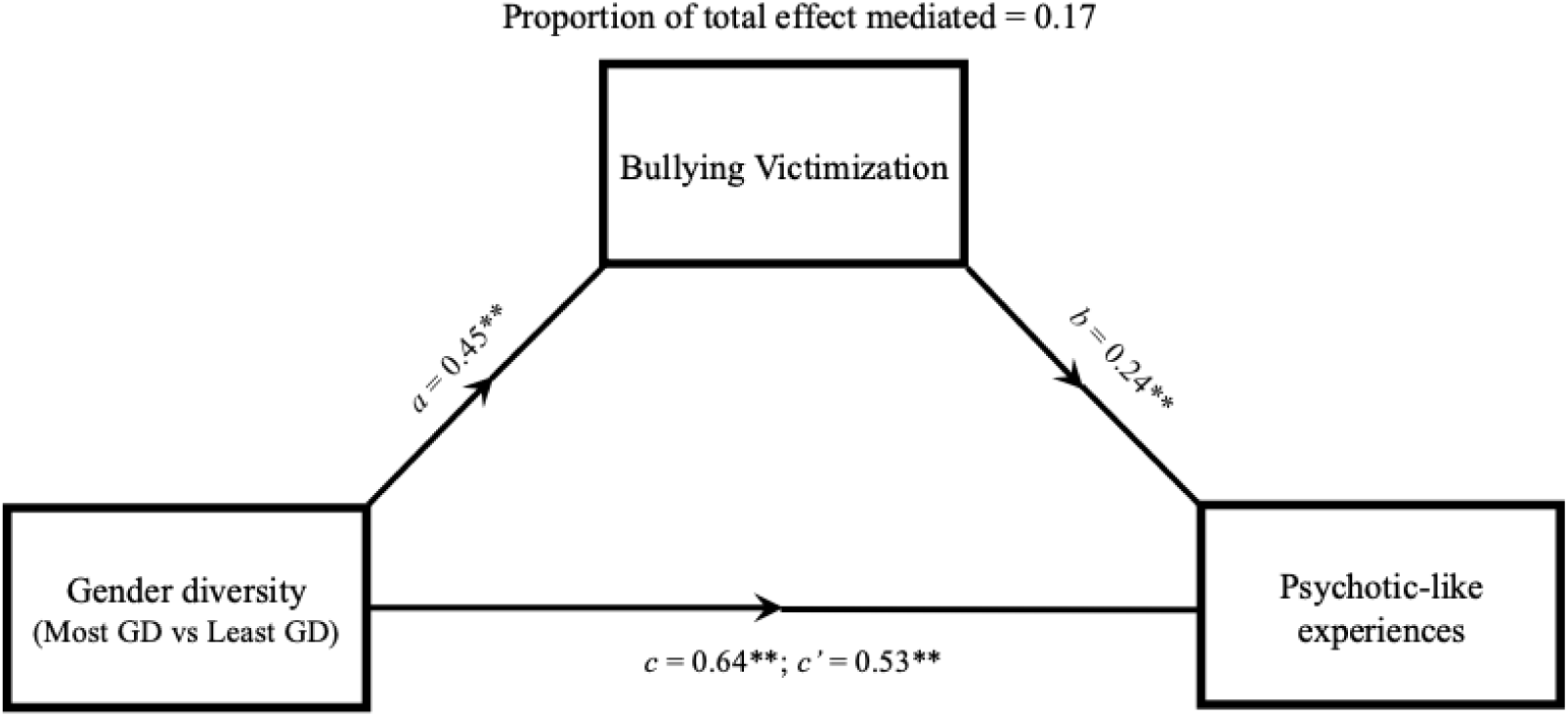
Path diagram representing the mediating effect of bullying victimization on the relationship between gender diversity and psychotic-like experiences (PQ-BC). Each model contained age, birth-assigned sex, pubertal development, parental education, family income, and site as fixed effects, and family ID as a random intercept; *c* = total effect, *c’* = direct effect, ** = p < 0.005; GD = gender diverse.

### Differential effects of bullying victimization on PLEs

Across all participants, bullying victimization was associated with significantly greater PLE scores (ß = 0.26, 95% CI [0.24, 0.27], p < 1×10^-16^; **Supplementary Tables 8, 9**). The effect of bullying victimization on PLEs was significantly stronger in the most gender diverse group compared to that in the least gender diverse group (ß = 0.14, 95% CI [0.09, 0.20], p = 5.1×10^-08^; see **Figure 3, Supplementary Table 10**).

**Figure 3.**
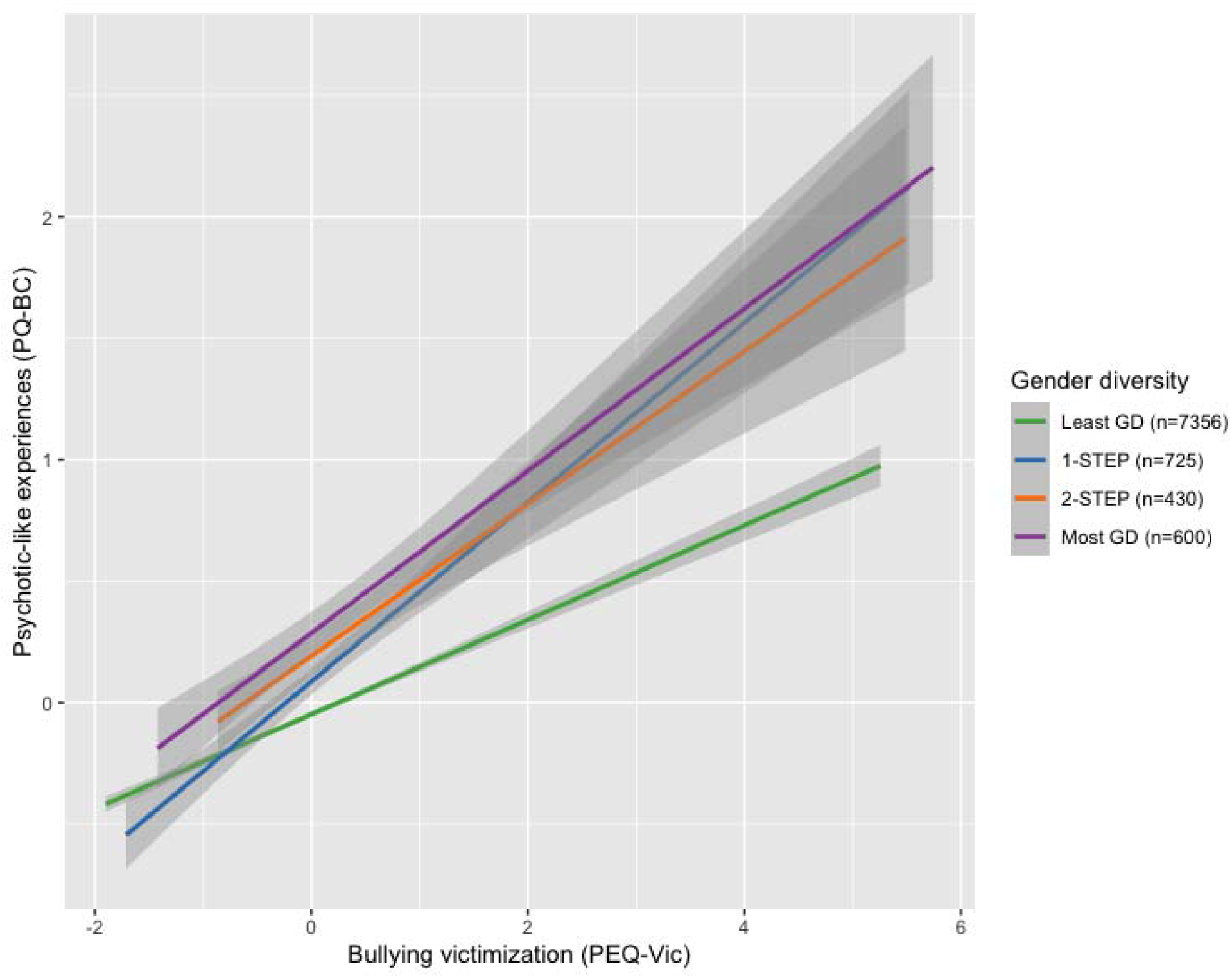
Relationship between bullying victimization (PEQ) and psychotic-like experiences (PQ-BC) by gender diversity group. Plotted values are residuals from models regressing out fixed effects covariates (age, birth-assigned sex, pubertal development, parental education, and family income), nesting family within site. Size of each analyzed gender diversity group is shown in parentheses next to group names.

Conversely, although greater bullying victimization was associated with higher broad mental health problem scores across all participants (ß=0.39, 95% CI [0.36, 0.41], p < 1 ×10^-16^; **Supplementary Table 11**), there was no interaction effect with gender diversity (ß = 0.06, 95% CI [-0.01, 0.14], p = 0.08; **Supplementary Table 12**), indicating similar effects of bullying on general psychopathology regardless of gender diversity.

### Comparison of cross-sectional associations in states with high and low support policies

Neither bullying victimization nor PLEs differed by gender diversity group in states with high versus low supportive gender identity-related laws at a single timepoint (**Supplementary Table 13**). The effect of bullying victimization on PLEs in the most gender-diverse group, however, differed as a function of state policies: in high (vs. low) support states, bullying victimization showed a weaker relationship with PLEs in the most gender diverse group (three-way interaction ß=-0.27, 95% CI [-0.42, -0.12], p=4.1×10^-4^; **Supplementary Table 14**). However, in post-hoc analyses in the most gender diverse group only, there was no difference in the effect of bullying victimization on PLEs in high versus low support states (**Supplementary Table 15**).

### Linear changes in psychotic-like experiences over time and relationship to state-level policies

Across Years 1 through 4, corresponding to average ages of 10.9 through 14.1 years, PLE scores declined by 0.10 standard deviations per year (ß = -0.10, 95% CI = [-0.13, -0.08], p = 2.2×10^-8^; **Supplementary Table 16**); that is, PLEs were less frequently endorsed over time in the whole sample. This longitudinal change in PLEs varied by gender diverse group such that, compared to the least gender diverse group, PLEs declined more slowly in the most gender diverse group (ß = 0.06, 95% CI [0.02, 0.10], p = 0.002; **Supplementary Table 17**). In states that consistently lacked supportive gender identity policies, PLEs *increased* over time in the most gender diverse youth, relative to least gender diverse youth residing in consistently supportive states (three-way interaction ß = 0.32, 95% CI [0.21, 0.43], p = 2.2×10^-8^; see **Figure 4, Supplementary Table 18, Supplementary Figure 2**). Post-hoc analyses revealed that within the most gender diverse group, PLEs increased over time in consistently low support states compared to consistently high support states (see **Supplemental Results**, **Supplementary Tables 19**). This three-way interaction effect was specific to PLEs and did not generalize to broad psychopathology, as measured by BPM (ß = 0.07, 95% CI [-0.06, 0.20], p = 0.28).

**Figure 4.**
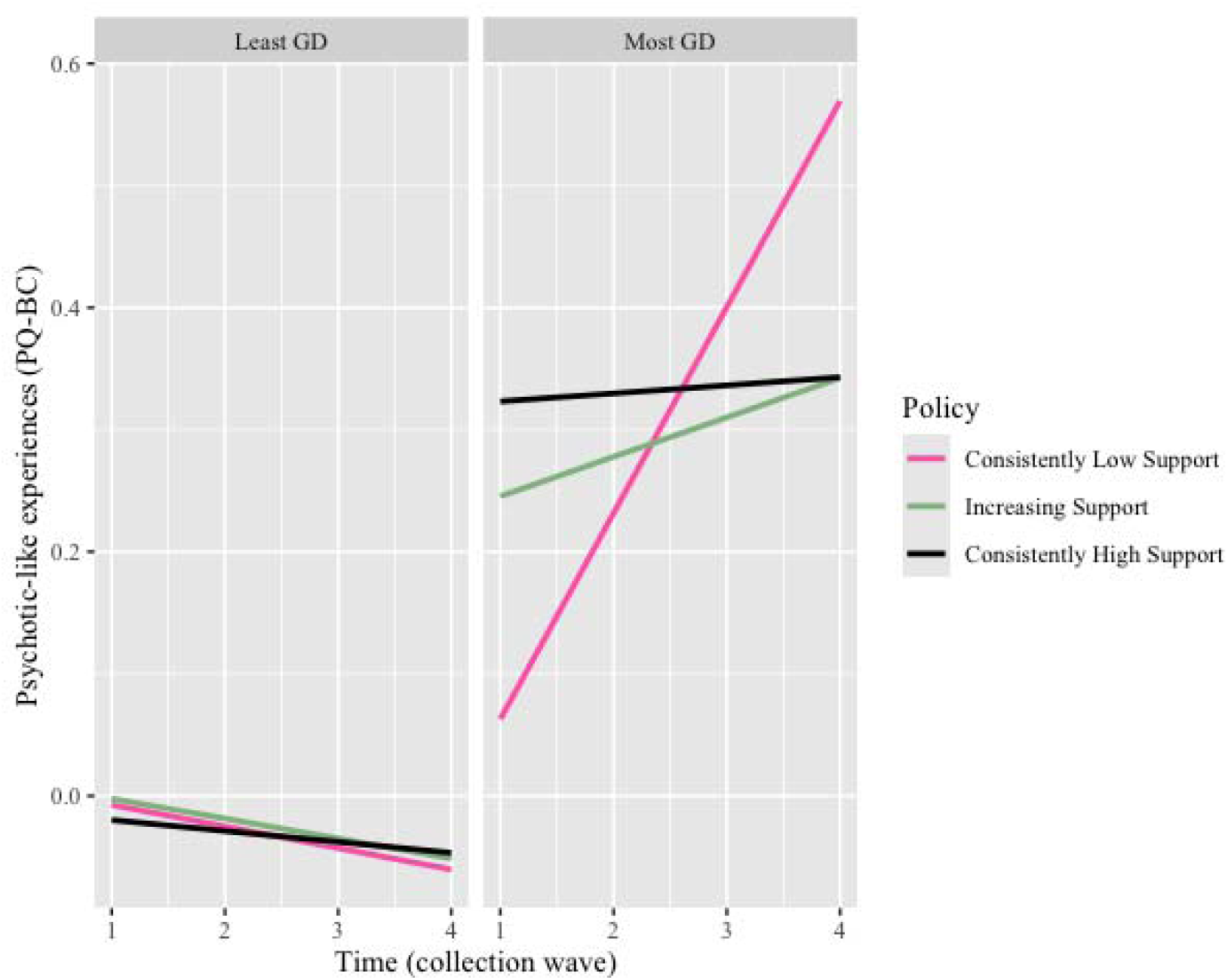
Representation of three-way interaction between time, state-level policy, and gender diversity. Psychotic-like experiences (PQ-BC) values represent residuals from models including covariates (birth-assigned sex, puberty levels, parental education, and family income), with slope over time allowed to vary within subjects nested within family within site. A regression line with a positive slope is interpreted as an increase in PQ-BC over time. The left panel represents data from the least gender diverse group, the right panel from the most gender diverse group. Pink lines plot data from states with consistently low policy-level support for gender identity, black with consistently high support, and green with increasing support over time. For reference, in the three-way interaction model, the slope of each of these lines is compared statistically to the black line in the left panel (i.e., PQ-BC change over time in the least gender diverse group in consistently supportive states). Only the least gender diverse group in states with consistently unsupportive policies shows a significantly different change (increase) in PQ-BC scores over time.

## DISCUSSION

Our analysis of selected distal stressors that contribute to self-reported psychotic-like experiences (PLEs) in youth recruited across 17 states, both at a single timepoint and over time, revealed several key findings with implications for clinical practice and policymaking. First, self-reported experiences of bullying victimization both mediated and moderated the relationship between gender diversity and PLEs. That is, more gender-diverse youth endorsed more PLEs, which was partially explained by increased reports of bullying; and the most gender diverse youth were more vulnerable to the effects of bullying victimization on PLEs. Second, unsupportive state policies exhibited nuanced effects on bullying victimization and PLEs: while there were no detectable effects of state policy at a single timepoint, more gender-diverse youth exhibited increases in PLEs over time in states that consistently lacked supportive legislation related to gender identity. In other states, regardless of gender diversity, PLEs declined or showed no change.

Our findings highlight the importance of considering both the local and state-level social environment when delivering care to gender-diverse youth. Recapitulating the importance of the social environment, another study using cross-sectional ABCD data found that stressful school environments contributed to broad mental health problems in gender diverse adolescents^33^. Anti-bullying interventions in schools have been shown to be cost-effective^34^, reduce bullying, and improve mental health among students^35^. PLEs in youth are a risk factor for the development of chronic psychotic disorders like schizophrenia^36^, which has an estimated annual economic burden of $343.2 billion in the U.S.^37^. Furthermore, PLEs index risk for poor functional outcomes and mental health problems, including suicidal behaviors^38,39^. Thus, anti-bullying measures may help promote well-being and prevent the onset of serious mental illness in gender-diverse young people. Additionally, given the role of interpersonal stressors in the occurrence of PLEs in gender diverse youth and the potential for misdiagnosis of psychosis^8^, it is critical for clinicians to consider minority stress in assessment and treatment planning^10^.

Findings suggest that chronic exposure to unsupportive legislation may exert negative effects on PLEs over time in gender diverse youth. The cross-sectional findings indicating no differences by policy in PLEs, bullying victimization, nor their relationship, were somewhat unexpected. Prior cross-sectional studies among TGD adults have shown that awareness of local anti-transgender laws was associated with mental health problems^40,41^ and that TGD adults in areas lacking protective laws reported more frequent exposure to discriminatory experiences^42^. The current study differs notably in that the sample consisted of adolescents who did not necessarily self-identify as TGD. Additionally, adolescents may have less awareness of state-level policy compared to TGD adults. Despite unexpected cross-sectional findings, longitudinal findings were aligned with our hypotheses and the minority stress framework, which characterizes minority stress as chronic^12^. Indeed, a recent study showed that the passing of anti-transgender legislation had the largest effects on suicidal behaviors 1 to 2 years later^20^. The current data show that in states with consistently unsupportive policies, PLEs increase in more gender diverse teens; whereas in all other groups PLEs declined or remained stable. Importantly, temporal changes in bullying victimization did not differ by state-level policy, suggesting that policy-associated stigma influences PLEs via mechanisms separate from bullying.

Interestingly, our findings revealed somewhat specific effects of minority stress on psychotic-like experiences in more gender diverse youth; the moderating effects of gender diversity did not generalize to broad mental health problems. Importantly, these findings do not refute ample, prior evidence that discrimination and interpersonal stressors contribute to other mental health problems like depression, anxiety, and suicidal ideation^6,13,43,44^. Rather they suggest that unsupportive policies have particularly strong and negative effects on PLEs. In other minoritized groups, stigma has been associated with increased hypervigilance^45^ and clinically significant paranoia^23^, a central^22^ and perpetuating^46^ feature of psychosis. Thus, such behaviors may arise in response to bullying victimization and policy-associated stigma and thus increase susceptibility of gender diverse youth to PLEs. These findings thus emphasize the importance of informed and sensitive assessment of such symptoms in this population.

### Limitations

Several limitations of the current study should be considered. As mentioned above, the nature of the policy measurement used precludes the investigation of the effects of specific policies. Additionally, the state-level policy variables provided by MAP represent the legislative climate at the start of each year in which a participant’s data were collected (January 1st). This timing limited the ability to assess specific temporal dynamics of policy effects on mental health. Furthermore, while state-level income inequality was controlled for, there may be other state-level characteristics that are correlated with gender identity-related legislation and influence. Additionally, participants’ states of residence were inferred from data collection sites rather than residential addresses, which are not publicly available.

## CONCLUSIONS

This study adds to a sparse literature on the effects of state-level policy on psychotic-like experiences in gender diverse adolescents, using a dimensional measure of gender independent of gender identity. The results suggest that: (1) regardless of self-endorsed gender identity, the mental health of youth with dimensionally incongruent experiences of gender is negatively impacted via mechanisms similar to those impacting TGD-identifying individuals; (2) psychotic-like experiences in the context of gender diversity are exacerbated by bullying; and, (3) gender diversity is only associated with the progression of psychotic-like experiences in the context of consistently unsupportive political environments. These data suggest that continued sociopolitical stigma against gender diversity may contribute to higher rates of psychotic-like experiences in youth.

## Data Availability

All ABCD data are available to investigators with valid Data Use Agreements here: https://www.nbdc-datahub.org/. State-level policy data can be requested from the Movement Advancement Project

## Acknowledgements

The ABCD Study is supported by the National Institutes of Health and additional federal partners via the following awards: U01DA041048, U01DA050989, U01DA051016, U01DA041022, U01DA051018, U01DA051037, U01DA050987, U01DA041174, U01DA041106, U01DA041117, U01DA041028, U01DA041134, U01DA050988, U01DA051039, U01DA041156, U01DA041025, U01DA041120, U01DA051038, U01DA041148, U01DA041093, U01DA041089, U24DA041123 and U24DA041147. A full list of federal supporters is available at https://abcdstudy.org/federal-partners.html. Participating study sites and site principal investigators can be found at https://abcdstudy.org/consortium_members/. We thank the investigators and staff at the ABCD sites and coordinating centers as well as the study participants and their families for their essential contributions to this work. This manuscript reflects the views of the authors and may not reflect the opinions or views of the NIH or ABCD consortium investigators. DEH is supported by the National Institute of Mental Health (T32 NS048004). We thank the team at Movement Advancement Project for curating state-level data on policies related to LGBTQ equality and for providing historical data for this project.

