## Supplement for "Association between bullying, state-level policy and mental health symptoms in gender diverse youth"

*TABLE OF CONTENTS*

Supplementary Methods ………………………………………………………………………………	2

Supplementary Results ..………………………………………………………………………………. 6

Supplementary Figures ..………………………………………………………………………………	8

Supplementary Figure 1

Supplementary Figure 2.

Supplementary Figure 3.

Supplementary Figure 4.

Supplementary Figure 5.

Supplementary Tables ….……………………………………………………………………………….. 14

Supplementary Table 1

Supplementary Table 2

Supplementary Table 3

Supplementary Table 4

Supplementary Table 5

Supplementary Table 6

Supplementary Table 7

Supplementary Table 8

Supplementary Table 9

Supplementary Table 10

Supplementary Table 11

Supplementary Table 12

Supplementary Table 13

Supplementary Table 14

Supplementary Table 15

Supplementary Table 16

Supplementary Table 17

Supplementary Table 18

Supplementary Table 19

Supplementary Table 20

Supplementary Table 21

**Supplementary Methods**

**Datasets**

The current analysis used all complete data available at the time of writing, which consisted of data from 4 timepoints (Year 1 through Year 4 follow up) from the ABCD data Release 5.1, accessed November 18, 2023. Baseline data were not analyzed as the Felt-gender questionnaire used to measure gender diversity^1^ (described below) was introduced into the ABCD protocol starting in the year 1 follow up. The Movement Advancement Project (MAP) provided data on state-level measures of gender-identity-related legislation. MAP data have been used in similar analyses in transgender adults^2^.

**Gender diversity**

Gender diversity was assessed via the Gender Identity and Sexual Health (GISH) survey^1^. The GISH survey measures three constructs related to gender identity: felt-gender, gender nonconformity (gender expression), and gender contentedness^3^. In the current analysis, the two items assessing felt-gender were used, which assess the respondent’s experience of gender. Subjects were assigned to one of four groups based on their responses to two questions: “How much do you feel like a boy/girl?” and “How much do you feel like a girl/boy?” (**Figure 1**) with response options on a 5-point Likert scale ranging from “Not at all” to “Totally”. If the respondent was assigned male at birth, for example, they would respond to the first question (“How much do you feel like a boy?”) and then the second question (“How much do you feel like a girl?”). The four groups progressed from least gender diverse (Least GD) to most gender diverse (Most GD), with the former representing complete congruence of felt-gender with birth-assigned sex (e.g., a birth-assigned male responding that they felt ‘totally’ like a boy and ‘not at all’ like a girl) and the latter representing an experience of gender most divergent from birth-assigned sex. The intermediate categories are labeled 1-STEP and 2-STEP and represent stepwise degrees of variation away from congruence with birth-assigned sex (**Figure 1**). Data collection for these data began during the year 1 follow-up, and thus baseline data were excluded from analyses; for cross-sectional analyses, data from year 3 follow-up was used; for longitudinal analyses, data from years 1 through 4 were used.

**Policy scoring**

Tallies of each state’s policies related to gender identity were collated and provided by the Movement Advancement Project (MAP). According to the methodology outlined on MAP’s website (see https://www.lgbtmap.org/equality_maps/profile_state/MO), the enactment of a supportive law counts as 0.5 points or 1 point for that state (depending on the impact of the bill), whereas legislation of a negative bill counts as -0.5 points to -1 point. MAP tallies the number of policies and sums the total points for each state. That is, each time a bill is passed in a state throughout the year, the state’s policy score changes based on the valence (negative or positive) and the impact (see **Supplementary Table 21** for examples of laws that might receive a +0.5, +1, -0.5, and -1). For the data used in the current study, since ABCD data were collected across different years, the policy tally represents the total number of points that a state received on January 1st of the respective year. Three categories of state tallies were considered, all of which were derived and provided by MAP: an Overall score, which represents a measure of LGBTQ+ -supportive laws broadly; a Gender Identity tally, representing a tally of laws in support of gender-diverse individuals; and a Youth tally, which represents a tally of laws supporting LGBTQ+ youth. While each of these domains are overlapping and all likely impact gender diverse youth, we used the Gender Identity tally for analyses, as we were specifically interested in the effect of laws related to gender identity. This score comprises tallies of laws that “explicitly address or impact gender identity and/or expression”.

***Calculation of policy scores***

Between years, the total number of possible points differs as new policies are enacted: in 2017, the max policy score was 16.5; in 2022, the max policy score was 21.75. As such, a comparison of raw policy scores was not appropriate, and thus raw scores were converted to a proportional score, whereby a score of 1 represented a state that had enacted all possible supportive laws in that year; a score of 0 represented a state that had not enacted any supportive laws, and/or had passed negative laws. Notably, some states had negative scores from MAP and thus their ‘proportion’ scores were negative. To illustrate the calculation of proportional scores: if the maximum tally for a given year across the United States was 10, and a particular state had received a tally of 6, the proportional tally score for that state would be 0.6.

For longitudinal analyses, as most state scores varied minimally across the 4 years, states were grouped into one of three categories: (1) states that scored consistently high (i.e., at or above 0.5; H-H), (2) consistently low (below 0.5; L-L), or (3) that increased by more than 0.23 proportional points across 4 years. A threshold of 0.23 was determined by the upper quantile of the difference scores, calculated by subtracting Year 4 scores from Year 1 scores. Notably, the distribution of difference scores was largely positive (**Supplementary Figure 3**); many of the negative values (indicating a decline in the gender identity tally proportion over time) were not meaningful/interpretable. For example, of the 1,123 subjects whose difference scores fell below the lower quartile cut-off (-0.006), 620 resided in California (CA) for both time points, where at Year 1 the mean proportional tally score was 0.97 and at Year 4 was 0.96. Likewise, the mean proportional tally score in Oklahoma (OK) at Year 1 was -0.13 and at Year 4, -0.14. While the *difference scores* of Oklahoma and California were both roughly equally negative, categorizing both of these states in the same group (i.e., “Decreasing”) would not be meaningful. There were 24 participants whose difference scores were more negative than those described above, due to their moving sites between Year 1 and Year 4. While these data may provide valuable insight into the effects of declining political support, due to the small sample size (n=24) and the unmeasured confound of stress related to moving, these participants were removed from analysis. Participants whose difference scores were negative and who participated at the same site at Year 1 and Year 4 (i.e., CA or OK residents) were classified in the stable category (i.e., either consistently high or consistently high), depending on whether the proportional tally scores at both time points were above or below 0.5.

Notably, the year of data collection varied within site and timepoint; that is, data collected at the same time point, at the same site/state, may have been collected in different calendar years. As a result of this, a participant’s value for the policy variable may differ from another participant from the same state depending on the year of data collection (**Supplementary Figure 4**). See **Supplementary Figure 5** for a visual of how state-level policy scores changed between 2017 and 2022.

**Prodromal Questionnaire - Brief Child Version (PQ-BC)**

The PQ-BC is a modified version of the Prodromal Questionnaire - Brief, which was originally designed and validated as a 21-item questionnaire used to screen individuals for psychotic spectrum syndromes^4^. The PQ-BC was designed and validated in ABCD to assess psychotic-like experiences by editing the language of the questions and adding picture anchors to be more easily comprehended by youth^5^. Subjects indicated whether they experienced each item (yes/no). If participants endorsed a given PLE, then they indicated whether the experience bothered them (yes/no), and if so, then subjects indicated the level of distress from the experience on a 5-point Likert scale (1 = not distressing, 5 = very distressing) with picture anchors to assist comprehension (see ref^5^ for more details). Cross-sectional analyses were performed on data from year 3 follow-up; for longitudinal analyses, data from years 1 through 4 were used.

**Brief Problem Monitor**

The Brief Problem Monitor (BPM) is an abbreviated version of the parent-reported Child Behavior Checklist that was completed by ABCD participants beginning at the second time point (Year 1). The BPM total score was analyzed, and not the CBCL total score, to measure broad mental health problems as assessed comparably to PLEs (i.e., via self-report). Cross-sectional analyses were performed on data from year 3 follow-up; for longitudinal analyses, data from years 1 through 4 were used.

**Covariates**

***Puberty***

Pubertal development stage, measured via a 5-item questionnaire completed by adolescents and parents^6^, was included as a covariate, as the development of gender identity is intertwined with the development of sex characteristics and puberty^7^. The Pubertal Development Scale measures parents’ report of their child’s stage of pubertal development based on reference to 5 specific physical characteristics (e.g., body hair growth, development of pimples, etc.)^6^. Parents answered each question on a 4-point Likert scale from “[development of physical characteristic] has not yet begun” to “seems complete”. The average of answers to all 5 questions was used in analysis as per recommendations and previous literature^8,9^. For longitudinal analyses (using data from years 1 through 4), the average of the 5 responses was calculated each year and included in the model as a time-varying fixed effect covariate.

***Race/Ethnicity***

Self-reported race and ethnicity were omitted from main analyses, though were included in supplementary models exploring the possible role of these social constructs as effect modifiers, although they were not the focus of the study (**Supplementary Tables 3, 6, 9**). Interpretation of effect sizes and associated statistics did not differ with or without the inclusion of race and ethnicity. Previous research has reported differences in rates of both PLEs and gender diversity between racial/ethnic groups^10,11^; thus, secondary analyses were conducted to test whether the reported primary effects of bullying victimization or policy on PLEs were modified by race/ethnicity. Because the effects of interest changed minimally, we opted to primarily report models without terms for race or ethnicity to avoid the propagation of ‘race differences’ when race was not central to our research question^12,13^. Notably, it is important for future research to probe the effects of policy and bullying victimization on mental health in groups with intersecting identities (i.e., groups with multiply-marginalized identities).

For supplementary analyses, the race variable was coded according to guidance from ABCD (https://github.com/ABCD-STUDY/analysis-nda) and comprised 6 levels: White, Black, Asian, American Indian/Alaskan Native/Native Hawaiian/Pacific Islander, Other, and Multiracial (more than one race). The variable for ethnicity was binary: Non-Hispanic/Latino/x and Hispanic/Latino/x.

***Gini Coefficient***

Supplementary models of analyses examining the effect of policy included each state’s Gini coefficient, a measure of state-level income inequality^14^. The Gini coefficient has been widely used and accepted as a gold standard for measuring income inequality. A value of 0 represents perfect equality, whereas a value of 1 represents perfect inequality. Gini coefficients for all U.S. states at each year represented in the ABCD sample (2018-2022) were collected as part of the American Community Survey and downloaded directly from the U.S. Census site (data.census.gov).

**Supplemental Results**

Post-hoc analyses were conducted to test the robustness of the interpretation that adolescents in the most gender diverse (Most GD) group from states with consistently unsupportive gender identity-related policy exhibit increases in PQ-BC over time, relative to other adolescents in the United States. Firstly, the interaction between time and longitudinal state policy ($\beta_{time x state policy}$) was tested in the most gender diverse group. This interaction effect was significant suggesting that the change in PQ-BC over time differed in the Most GD participants in consistently unsupportive states compared to Most GD participants in consistently supportive states (ß=0.30, p=0.006, 95% CI [0.09,0.51]; **Supplementary Table 19**). Next, to probe the initial 3-way interaction term further, we stratified the groups by gender diversity group (k=4) and longitudinal state law (k=3), and regressed PLEs (residualized for birth-assigned sex, pubertal level, race/ethnicity, parental education, and family income) on time in each group, correcting for 12 comparisons. Adolescents in the Most GD group within LL states were the only group that exhibited a significant increase over time in residualized PQ-BC scores after adjusting for multiple comparisons (ß = 0.18, 95% CI [0.06, 0.31], p = 3.42x10^-3^, q < 0.05; **Supplementary Table 20**).

**Supplementary Figures**

**Supplementary Figure 1.**

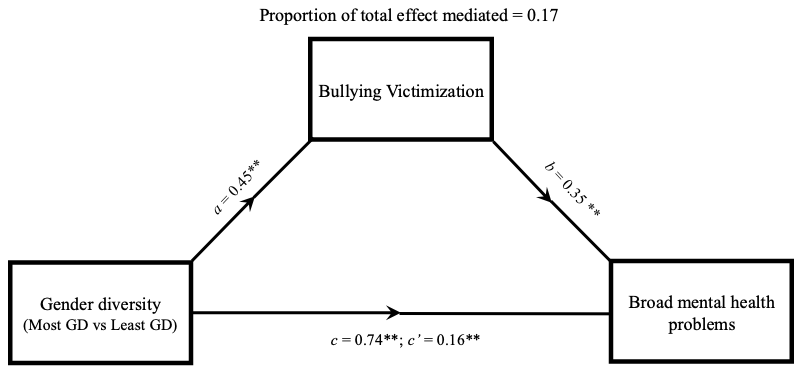

**Supplementary Figure 1**. Path diagram representing the mediating effect of bullying victimization on the relationship between gender diversity and broad mental health problems (BPM total). Each model contained age, birth-assigned sex, pubertal development, parental education, family income, and site as fixed effects, and family ID as a random intercept. Most GD = most gender-diverse, Least GD = least gender diverse; *c* = total effect, *c’* = direct effect, ** = p < 0.005.

**Supplementary Figure 2**.

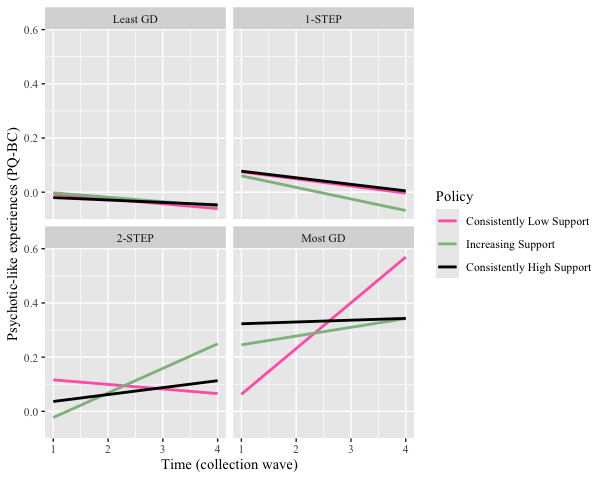

**Supplementary Figure 2.** Representation of three-way interaction between time, state-level policy, and gender diversity group. This figure differs from **Figure 3** (main text) in that it includes the 1-STEP and 2-STEP gender diversity groups. PQ-BC values represent PQ-BC residuals from models including covariates (birth-assigned sex, puberty levels, parental education, and family income), with slope over time allowed to vary within subjects nested within family within site. A regression line with a positive slope can be interpreted as an increase in PQ-BC over time. Each panel represents data from each of the 4 gender diversity groups. Pink lines plot data from states with consistently low political support for gender identity, black with consistently high support, and green with increasing support over time. For reference, in the three-way interaction model, the slope of each of these lines is compared statistically to the black line in the top left panel (i.e., PQ-BC change over time in the least gender diverse group in consistently supportive states). Only the most gender diverse group in consistently unsupportive states shows a significantly different change (increase) in PQ-BC over time.

**Supplemental Figure 3**

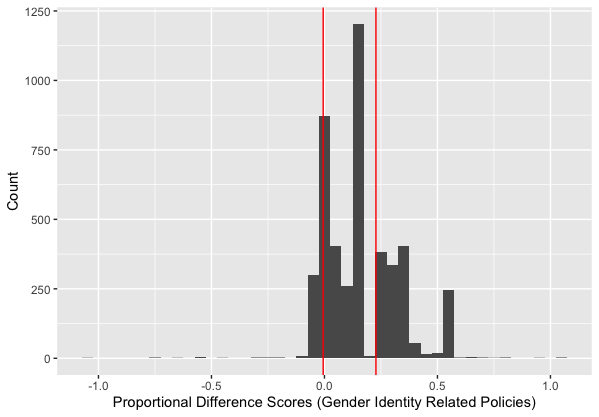

**Supplemental Figure 3.** Distribution of policy proportional difference scores. The difference score was calculated by subtracting the Year 1 from Year 4 policy proportion scores (i.e., policy score for a state divided by the max policy score across states for a given year, as provided by MAP). The vertical red lines indicate quartile cut-offs. Values between the cutoffs represent participants within states that showed relatively no change in policy tallies across four years. Right-most values represent participants within states that showed increases in policy tallies, indicating increases in supportive policy over time; left-most values represent those that showed decreases in policy tallies, indicating decreases in supportive policy over time.

**Supplementary Figure 4.**

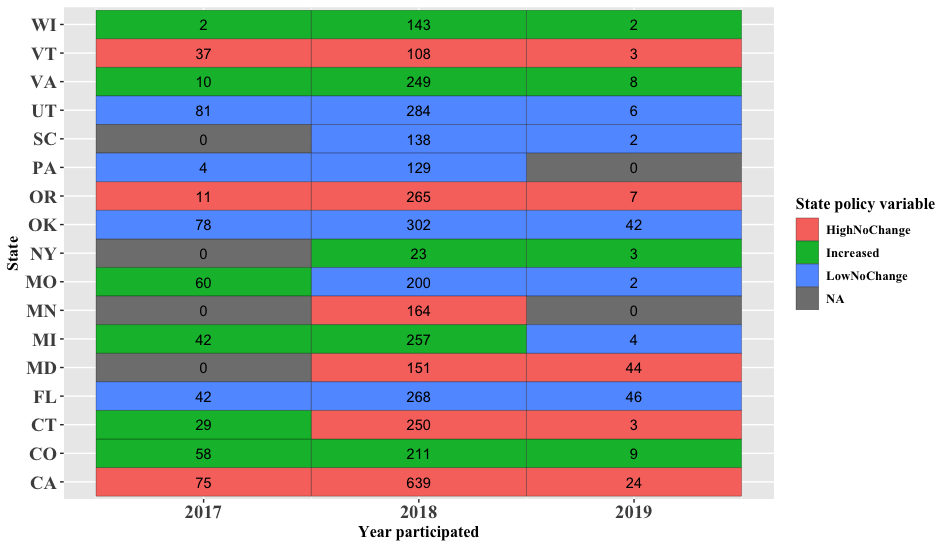

**Supplemental Figure 4.** Depiction of the influence of participation year on the derivation of the state policy variable. Numbers represent the size of each group. Because the longitudinal state-level policy variable (i.e., High, No Change; Increased; Low No Change) was derived per individual based on their interview date, individuals within the same state may not be assigned to the same state-level policy category. For example, individuals whose Year 1 data were collected in 2017 and who participated at the Missouri site (Washington University at St. Louis) were categorized in the “Increased” group; however, participants enrolled in the same site whose data were collected in 2018 or 2019 were categorized in the “Low No Change” group. That is, there were 60 individuals who participated in the study in 2017 in Missouri for whom at the time of their Year 4 data collection, the state exhibited an increase in gender identity related policy tally as measured by MAP. Likewise, the categorization of participants into policy change levels was influenced by the year in which Year 4 data were collected for a given participant.

**Supplemental Figure 5.**

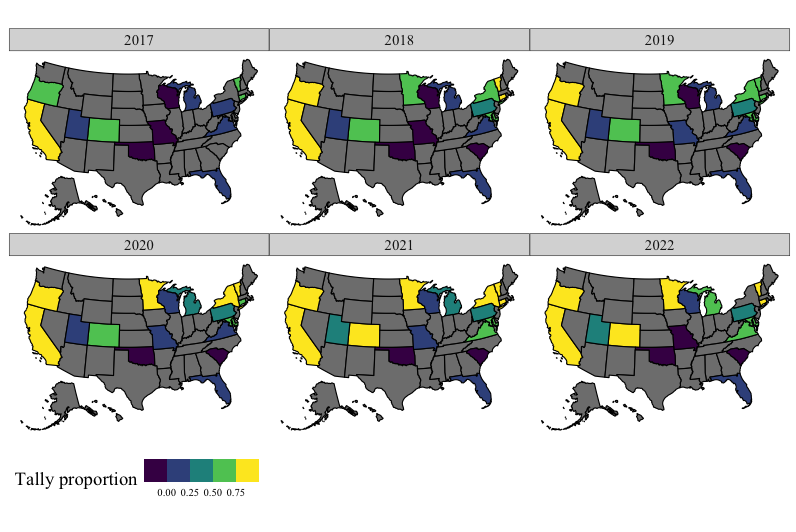

**Supplemental Figure 5.** Map of the United States showing the Gender Identity Tally proportion variable across time in the ABCD sample. A value of 1 indicates a state that had enacted all possible positive laws in that year (shown above each map), whereas a value of 0 would indicate a state that had enacted no positive laws. A negative value indicates a state where the sum of policy tallies (according to MAP’s definition) was negative. Thus, states towards the yellow end of the color spectrum are considered to have more supportive policies related to gender identity; those toward the indigo end of the spectrum have relatively unsupportive policies. Grey states represent states in which data were not collected in ABCD or data were not available.

**Supplementary Tables**

**Supplementary Table 1**

| **Gender Identity** | **Least GD (n=8,240)** | **1-STEP (n=825)** | **2-STEP (n=491)** | **Most GD (n=689)** |
| --- | --- | --- | --- | --- |
| Female | 3282 (40.6) | 531 (66.5) | 376 (79.8) | 436 (67.9) |
| Male | 4778 (59.2) | 260 (32.6) | 92 (19.5) | 107 (16.7) |
| Trans Female | 3 (0.0) | 0 (0.0) | 0 (0.0) | 9 (1.4) |
| Trans Male | 2 (0.0) | 2 (0.3) | 0 (0.0) | 24 (3.7) |
| Gender Queer | 7 (0.1) | 4 (0.5) | 1 (0.2) | 39 (6.1) |
| Different | 3 (0.0) | 1 (0.1) | 2 (0.4) | 27 (4.2) |

**Supplementary Table 1. Gender diversity groupings are partially overlapping with, yet independent from, gender identity.** Breakdown of participant-reported gender identity stratified by gender diversity groups (Least GD, 1-STEP, 2-STEP, Most GD). In the ABCD study, participants could report their gender identity as female, male, trans female, trans male, gender queer, or different. Although most participants that identified as trans, genderqueer, or different were included in the most gender diverse group, some were not. Likewise, some identifying as male or female (cis) were included in the most gender diverse group, indicating that experience of gender and gender identity are related but measure slightly different constructs.

**Supplementary Table 2**

| **Term (DV = PQBC)** | **Estimate** | **Std.Error** | **P** | **95% CI** |
| --- | --- | --- | --- | --- |
| (Intercept) | 1.218 | 0.1959 | 5.35e-10 | [0.834, 1.602] |
| Gender diversity (1-STEP) | 0.248 | 0.0314 | 3.23e-15 | [0.187, 0.31] |
| Gender diversity (2-STEP) | 0.472 | 0.0406 | 5.62e-31 | [0.392, 0.551] |
| **Gender diversity (Most GD)** | **0.633** | **0.0354** | **3.23e-70** | **[0.564, 0.703]** |
| Age | -0.068 | 0.0144 | 2.49e-06 | [-0.096, -0.04] |
| Birth-assigned sex (Male) | 0.003 | 0.0211 | 0.894 | [-0.039, 0.044] |
| Puberty | 0.042 | 0.011 | 1.2e-04 | [0.021, 0.064] |
| Parental education | -0.022 | 0.0048 | 6.22e-06 | [-0.031, -0.012] |
| Family income | -0.02 | 0.0052 | 1.96e-04 | [-0.03, -0.009] |

**Supplementary Table 2. Participants in the most gender diverse group report greater psychotic-like experiences (PQ-BC).** Statistics from cross-sectional, linear mixed effects model regressing PQ-BC (psychotic-like experiences; dependent variable) on gender diversity groups and covariates (fixed effects) allowing a random intercept by family within site. Effects of primary interest in bold. Referent groups: gender diversity (Least GD), birth-assigned sex (female). Units: age (years); puberty (scaled; mean = 0, standard deviation = 1); parental education (number of education years completed); family income, 10 levels ranging from 1 (< $5,000 yearly) to 10 (> $200,000 yearly).

**Supplementary Table 3.**

| **Term (DV = PQ-BC)** | **Estimate** | **Std.Error** | **P** | **95% CI** |
| --- | --- | --- | --- | --- |
| (Intercept) | 0.985 | 0.2013 | 1.02e-06 | [0.59, 1.379] |
| Gender diversity (1-STEP) | 0.253 | 0.0315 | 1.11e-15 | [0.191, 0.315] |
| Gender diversity (2-STEP) | 0.477 | 0.0407 | 1.52e-31 | [0.398, 0.557] |
| **Gender diversity (Most GD)** | **0.639** | **0.0356** | **8.77e-71** | **[0.569, 0.709]** |
| Age | -0.062 | 0.0145 | 1.56e-05 | [-0.091, -0.034] |
| Birth-assigned sex (Male) | 0.004 | 0.0212 | 0.852 | [-0.038, 0.045] |
| Puberty | 0.036 | 0.0111 | 0.001 | [0.014, 0.058] |
| Parental education | -0.019 | 0.005 | 2.02e-04 | [-0.028, -0.009] |
| Family income | -0.011 | 0.0055 | 0.047 | [-0.022, 0] |
| Race (Black) | 0.171 | 0.0312 | 4.65e-08 | [0.11, 0.232] |
| Race (Asian) | -0.024 | 0.0612 | 0.697 | [-0.144, 0.096] |
| Race (AIAN/NHPI) | 0.278 | 0.1129 | 0.014 | [0.056, 0.499] |
| Race (Other) | 0.016 | 0.0502 | 0.756 | [-0.083, 0.114] |
| Race (Multiracial) | 0.088 | 0.0275 | 0.001 | [0.034, 0.142] |
| Ethnicity (Hispanic) | 0.053 | 0.0275 | 0.054 | [-0.001, 0.107] |

**Supplementary Table 3. Participants in the most gender diverse group report greater psychotic-like experiences (PQ-BC), with race and ethnicity as possible effect modifiers.** Statistics from cross-sectional linear mixed effects model regressing PQ-BC (psychotic-like experiences; dependent variable) on gender diversity groups and fixed-effects covariates including terms for race and ethnicity, allowing a random intercept by family within site. This table differs from Supplementary Table 2 only in that it includes terms for race and ethnicity. Effects of primary interest in bold. Referent groups: gender diversity (Least GD), birth-assigned sex (female), race (White), ethnicity (Non-hispanic). Units: age (years); puberty (scaled; mean = 0, standard deviation = 1); parental education (number of education years completed); family income, 10 levels ranging from 1 (< $5,000 yearly) to 10 (> $200,000 yearly). AIAN/HNPI = American Indian/Native Alaskan/Hawaiian/Native Pacific Islander

**Supplementary Table 4.**

| **Term (DV = BPM total)** | **Estimate** | **Std.Error** | **P** | **95% CI** |
| --- | --- | --- | --- | --- |
| (Intercept) | -0.315 | 0.2557 | 0.218 | [-0.816, 0.186] |
| Gender diversity (1-STEP) | 0.445 | 0.0415 | 1e-26 | [0.364, 0.527] |
| Gender diversity (2-STEP) | 0.833 | 0.0533 | 3.86e-54 | [0.728, 0.938] |
| **Gender diversity (Most GD)** | **0.896** | **0.0476** | **2.85e-77** | **[0.803, 0.989]** |
| Age | 0.046 | 0.0188 | 0.013 | [0.01, 0.083] |
| Birth-assigned sex (Male) | 0.061 | 0.0274 | 0.026 | [0.007, 0.115] |
| Puberty | 0.05 | 0.0143 | 5.24e-04 | [0.022, 0.078] |
| Parental education | -0.017 | 0.0064 | 0.008 | [-0.029, -0.004] |
| Family income | -0.013 | 0.0069 | 0.068 | [-0.026, 0.001] |

**Supplementary Table 4. Participants in the most gender diverse group report greater broad mental health problems (BPM total).** Statistics from cross-sectional, linear mixed effects model regressing BPM total (broad mental health problems; dependent variable) on gender diversity groups and covariates (fixed effects) allowing a random intercept by family within site. Effects of primary interest in bold. Referent groups: gender diversity (Least GD), birth-assigned sex (female). Units: age (years); puberty (scaled; mean = 0, standard deviation = 1); parental education (number of education years completed); family income, 10 levels ranging from 1 (< $5,000 yearly) to 10 (> $200,000 yearly).

**Supplementary Table 5.**

| **Term (DV = PEQ-Vic)** | **Estimate** | **Std.Error** | **P** | **95% CI** |
| --- | --- | --- | --- | --- |
| (Intercept) | -0.77 | 0.2425 | 0.001 | [-1.246, -0.295] |
| Gender diversity (1-STEP) | 0.151 | 0.039 | 1.06e-04 | [0.075, 0.227] |
| Gender diversity (2-STEP) | 0.404 | 0.0504 | 1.18e-15 | [0.305, 0.503] |
| **Gender diversity (Most GD)** | **0.476** | **0.0439** | **2.87e-27** | **[0.39, 0.563]** |
| Age | 0.05 | 0.0178 | 0.005 | [0.015, 0.085] |
| Birth-assigned sex (Male) | 0.041 | 0.0261 | 0.113 | [-0.01, 0.093] |
| Puberty | 0.013 | 0.0136 | 0.352 | [-0.014, 0.039] |
| Parental education | 0.001 | 0.006 | 0.878 | [-0.011, 0.013] |
| Family income | 0 | 0.0065 | 0.992 | [-0.013, 0.013] |

**Supplementary Table 5. Participants in the most gender diverse group report more frequent experiences of bullying victimization (PEQ-Victimization).** Statistics from cross-sectional, linear mixed effects model regressing PEQ-Vic (bullying victimization; dependent variable) on gender diversity groups and covariates (fixed effects) allowing a random intercept by family within site. Referent groups: gender diversity (Least GD), birth-assigned sex (female). Effects of primary interest in bold. Units: age (years); puberty (scaled; mean = 0, standard deviation = 1); parental education (number of education years completed); family income, 10 levels ranging from 1 (< $5,000 yearly) to 10 (> $200,000 yearly).

**Supplementary Table 6.**

| **Term (DV = PEQ-Vic)** | **Estimate** | **Std.Error** | **P** | **95% CI** |
| --- | --- | --- | --- | --- |
| (Intercept) | -0.502 | 0.2506 | 0.045 | [-0.994, -0.011] |
| Gender diversity (1-STEP) | 0.157 | 0.0394 | 7.05e-05 | [0.079, 0.234] |
| Gender diversity (2-STEP) | 0.399 | 0.0508 | 4.49e-15 | [0.3, 0.499] |
| **Gender diversity (Most GD)** | **0.471** | **0.0444** | **4.14e-26** | **[0.384, 0.558]** |
| Age | 0.041 | 0.018 | 0.024 | [0.005, 0.076] |
| Birth-assigned sex (Male) | 0.052 | 0.0264 | 0.049 | [0, 0.104] |
| Puberty | 0.023 | 0.0138 | 0.092 | [-0.004, 0.05] |
| Parental education | -0.003 | 0.0062 | 0.579 | [-0.016, 0.009] |
| Family income | -0.004 | 0.0069 | 0.59 | [-0.017, 0.01] |
| Race (Black) | -0.045 | 0.0388 | 0.243 | [-0.121, 0.031] |
| Race (Asian) | -0.356 | 0.0761 | 2.97e-06 | [-0.505, -0.207] |
| Race (AIAN/NHPI) | 0.062 | 0.1407 | 0.661 | [-0.214, 0.337] |
| Race (Other) | -0.086 | 0.0624 | 0.166 | [-0.209, 0.036] |
| Race (Multiracial) | -0.032 | 0.0343 | 0.344 | [-0.1, 0.035] |
| Ethnicity (Hispanic) | -0.147 | 0.0339 | 1.46e-05 | [-0.214, -0.081] |

**Supplementary Table 6. Participants in the most gender diverse group report more frequent experiences of bullying victimization (PEQ-Victimization) with race and ethnicity as possible effect modifiers.** Statistics from linear mixed effects model regressing PEQ-Vic (bullying victimization; dependent variable) on gender diversity groups and fixed-effects covariates including terms for race and ethnicity allowing a random intercept by family within site. This table differs from Supplementary Table 4 only in that it includes terms for race and ethnicity. Effects of primary interest in bold. Referent groups: gender diversity (Least GD), birth-assigned sex (female), race (White), ethnicity (Non-Hispanic). Units: age (years); puberty (scaled; mean = 0, standard deviation = 1); parental education (number of education years completed); family income, 10 levels ranging from 1 (< $5,000 yearly) to 10 (> $200,000 yearly).

**Supplementary Table 7.**

| **Term (DV = PEQ-Vic)** | **Estimate** | **Std.Error** | **P** | **95% CI** |
| --- | --- | --- | --- | --- |
| (Intercept) | -0.061 | 0.2052 | 0.765 | [-0.463, 0.341] |
| Gender diversity (1-STEP) | 0.052 | 0.0332 | 0.116 | [-0.013, 0.117] |
| Gender diversity (2-STEP) | 0.271 | 0.0429 | 2.83e-10 | [0.187, 0.355] |
| **Gender diversity (Most GD)** | **0.358** | **0.0373** | **1.29e-21** | **[0.284, 0.431]** |
| Age | 0.008 | 0.0151 | 0.615 | [-0.022, 0.037] |
| Birth-assigned sex (Male) | -0.077 | 0.0222 | 5.48e-04 | [-0.121, -0.033] |
| Puberty | 0.003 | 0.0115 | 0.828 | [-0.02, 0.025] |
| Parental education | -0.007 | 0.005 | 0.168 | [-0.017, 0.003] |
| Family income | 0.009 | 0.0055 | 0.098 | [-0.002, 0.02] |
| PEQ Perpetration | 0.546 | 0.0095 | <2e-16 | [0.527, 0.565] |

**Supplementary Table 7. Participants in the most gender diverse group report more frequent experiences of bullying victimization (PEQ-Victimization) after controlling for experiences of bullying perpetration (PEQ Perpetration).** Statistics from linear mixed effects model regressing PEQ-Vic (bullying victimization; dependent variable) on gender diversity groups and fixed-effects covariates including a term for PEQ-Perp (bullying perpetration) allowing a random intercept by family within site. Effects of primary interest in bold. Referent groups: gender diversity (Least GD), birth-assigned sex (female). Units: age (years); puberty (scaled; mean = 0, standard deviation = 1); parental education (number of education years completed); family income, 10 levels ranging from 1 (< $5,000 yearly) to 10 (> $200,000 yearly); PEQ Perpetration (scaled; mean = 0, standard deviation = 1).

**Supplementary Table 8.**

| **Term (DV = PEQ-Vic)** | **Estimate** | **Std.Error** | **P** | **95% CI** |
| --- | --- | --- | --- | --- |
| (Intercept) | 1.733 | 0.1899 | 8.99e-20 | [1.361, 2.105] |
| **PEQ Victimization** | **0.257** | **0.0084** | **7.3e-193** | **[0.24, 0.273]** |
| Age | -0.095 | 0.014 | 1.57e-11 | [-0.122, -0.067] |
| Birth-assigned sex (Male) | -0.077 | 0.0201 | 1.3e-04 | [-0.116, -0.038] |
| Puberty | 0.053 | 0.0107 | 6.69e-07 | [0.032, 0.074] |
| Parental education | -0.022 | 0.0047 | 1.74e-06 | [-0.031, -0.013] |
| Family income | -0.023 | 0.0051 | 5.97e-06 | [-0.033, -0.013] |

**Supplementary Table 8. Regardless of gender diversity, experiences of bullying victimization (PEQ Victimization) are associated with psychotic-like experiences (PQ-BC).** Statistics from cross-sectional, linear mixed effects model regressing PQ-BC (psychotic-like experiences; dependent variable) on PEQ-Vic (bullying) and fixed-effects covariates allowing a random intercept by family within site. Effects of primary interest in bold. Referent groups: birth-assigned sex (female). Units: PEQ Victimization (scaled; mean = 0, sd = 1); age (years); puberty (scaled; mean = 0, standard deviation = 1); parental education (number of education years completed); family income, 10 levels ranging from 1 (< $5,000 yearly) to 10 (> $200,000 yearly).

**Supplementary Table 9**

| **Term (DV = PEQ-Vic)** | **Estimate** | **Std.Error** | **P** | **95% CI** |
| --- | --- | --- | --- | --- |
| (Intercept) | 1.456 | 0.1951 | 9.31e-14 | [1.074, 1.838] |
| **PEQ Victimization** | **0.256** | **0.0085** | **2.58e-190** | **[0.24, 0.273]** |
| Age | -0.088 | 0.0141 | 4.52e-10 | [-0.115, -0.06] |
| Birth-assigned sex (Male) | -0.079 | 0.0202 | 8.75e-05 | [-0.119, -0.04] |
| Puberty | 0.044 | 0.0108 | 3.65e-05 | [0.023, 0.066] |
| Parental education | -0.019 | 0.0049 | 1.41e-04 | [-0.028, -0.009] |
| Family income | -0.014 | 0.0054 | 0.009 | [-0.025, -0.004] |
| Race (Black) | 0.172 | 0.0303 | 1.36e-08 | [0.113, 0.232] |
| Race (Asian) | 0.05 | 0.0595 | 0.402 | [-0.067, 0.167] |
| Race (AIAN/NHPI) | 0.272 | 0.1096 | 0.013 | [0.057, 0.487] |
| Race (Other) | 0.035 | 0.0488 | 0.479 | [-0.061, 0.13] |
| Race (Multiracial) | 0.113 | 0.0267 | 2.4e-05 | [0.06, 0.165] |
| Ethnicity (Hispanic) | 0.086 | 0.0267 | 0.001 | [0.033, 0.138] |

**Supplementary Table 9. Regardless of gender diversity, experiences of bullying victimization (PEQ Victimization) are associated with psychotic-like experiences (PQ-BC) with race and ethnicity as possible effect modifiers.** Statistics from linear mixed effects model regressing PQ-BC (psychotic-like experiences; dependent variable) on PEQ-Vic (bullying) and fixed-effects covariates, including terms for race and ethnicity, while allowing a random intercept by family within site. *This model differs from the model represented in Supplementary Table 8 in that it includes race and ethnicity as a fixed effect covariate.* Effects of primary interest in bold. Referent groups: birth-assigned sex (female), race (White), ethnicity (Non-Hispanic). Units: PEQ Victimization (scaled; mean = 0, sd = 1); age (years); puberty (scaled; mean = 0, standard deviation = 1); parental education (number of education years completed); family income, 10 levels ranging from 1 (< $5,000 yearly) to 10 (> $200,000 yearly).

**Supplementary Table 10.**

| **Term (DV = PQ-BC)** | **Estimate** | **Std.Error** | **P** | **95% CI** |
| --- | --- | --- | --- | --- |
| (Intercept) | 1.386 | 0.1868 | 1.3e-13 | [1.02, 1.753] |
| PEQ Victimization | 0.196 | 0.0098 | 2.88e-87 | [0.176, 0.215] |
| Gender diversity (1-STEP) | 0.206 | 0.0301 | 8.26e-12 | [0.147, 0.265] |
| Gender diversity (2-STEP) | 0.351 | 0.0399 | 1.9e-18 | [0.272, 0.429] |
| Gender diversity (Most GD) | 0.485 | 0.0351 | 5.19e-43 | [0.416, 0.554] |
| Age | -0.079 | 0.0137 | 1.04e-08 | [-0.106, -0.052] |
| Birth-assigned sex (Male) | -0.008 | 0.0201 | 0.676 | [-0.048, 0.031] |
| Puberty | 0.04 | 0.0105 | 1.54e-04 | [0.019, 0.06] |
| Parental education | -0.022 | 0.0046 | 1.45e-06 | [-0.031, -0.013] |
| Family income | -0.02 | 0.005 | 8.5e-05 | [-0.029, -0.01] |
| PEQ Victimization x Gender diversity (1-STEP) | 0.184 | 0.0306 | 1.95e-09 | [0.124, 0.244] |
| PEQ Victimization x Gender diversity (2-STEP) | 0.122 | 0.0314 | 1.11e-04 | [0.06, 0.183] |
| **PEQ Victimization x Gender diversity (Most GD)** | **0.144** | **0.0264** | **5.12e-08** | **[0.092, 0.195]** |

**Supplementary Table 10. The effect of bullying victimization (PEQ-Victimization) on psychotic-like experiences (PQ-BC) is larger in the most gender diverse compared to least gender diverse group (PEQ Victimization x Gender diversity (Most GD) term).** Statistics from cross-sectional, linear mixed effects model regressing PQ-BC (psychotic-like experiences; dependent variable) on PEQ-Victimization (bullying), gender diversity groups, and the interaction between the two. Effects of primary interest in bold. Fixed-effects covariates were age, sex, puberty, education, and family income. Random intercepts were allowed by family within site. Referent groups: gender diversity (Least GD); birth-assigned sex (female). Units: PEQ Victimization (scaled; mean = 0, sd = 1); age (years); puberty (scaled; mean = 0, standard deviation = 1); parental education (number of education years completed); family income, 10 levels ranging from 1 (< $5,000 yearly) to 10 (> $200,000 yearly).

**Supplementary Table 11.**

| **Term (DV = BPM total)** | **Estimate** | **Std.Error** | **P** | **95% CI** |
| --- | --- | --- | --- | --- |
| (Intercept) | 0.431 | 0.2457 | 0.08 | [-0.051, 0.912] |
| **PEQ Victimization** | **0.385** | **0.011** | **1.53e-248** | **[0.363, 0.407]** |
| Age | 0.009 | 0.0181 | 0.628 | [-0.027, 0.044] |
| Birth-assigned sex (Male) | -0.06 | 0.0259 | 0.02 | [-0.111, -0.01] |
| Puberty | 0.065 | 0.0138 | 2.56e-06 | [0.038, 0.092] |
| Parental education | -0.018 | 0.0061 | 0.003 | [-0.03, -0.006] |
| Family income | -0.017 | 0.0066 | 0.012 | [-0.029, -0.004] |

**Supplementary Table 11. Regardless of gender diversity, experiences of bullying victimization (PEQ Victimization) are associated with broad mental health problems (BPM total).** Statistics from linear mixed effects model regressing BPM total (broad mental health problems; dependent variable) on PEQ-Vic (bullying) and fixed-effects covariates allowing a random intercept by family within site. Effects of primary interest in bold. Referent groups: birth-assigned sex (female). Units: PEQ Victimization (scaled; mean = 0, sd = 1); age (years); puberty (scaled; mean = 0, standard deviation = 1); parental education (number of education years completed); family income, 10 levels ranging from 1 (< $5,000 yearly) to 10 (> $200,000 yearly).

**Supplementary Table 12.**

| **Term (DV = BPM total)** | **Estimate** | **Std.Error** | **P** | **95% CI** |
| --- | --- | --- | --- | --- |
| (Intercept) | -0.076 | 0.2394 | 0.751 | [-0.545, 0.393] |
| PEQ Victimization | 0.34 | 0.0125 | 1.57e-155 | [0.315, 0.364] |
| Gender diversity (1-STEP) | 0.384 | 0.0391 | 1.18e-22 | [0.308, 0.461] |
| Gender diversity (2-STEP) | 0.678 | 0.0517 | 5.79e-39 | [0.577, 0.78] |
| Gender diversity (Most GD) | 0.732 | 0.0462 | 7.92e-56 | [0.642, 0.823] |
| Age | 0.033 | 0.0176 | 0.064 | [-0.002, 0.067] |
| Birth-assigned sex (Male) | 0.045 | 0.0256 | 0.077 | [-0.005, 0.095] |
| Puberty | 0.044 | 0.0134 | 9.72e-04 | [0.018, 0.07] |
| Parental education | -0.018 | 0.0059 | 0.002 | [-0.03, -0.007] |
| Family income | -0.012 | 0.0064 | 0.067 | [-0.024, 0.001] |
| PEQ Victimization x Gender diversity (1-STEP) | 0.07 | 0.0391 | 0.073 | [-0.007, 0.147] |
| PEQ Victimization x Gender diversity (2-STEP) | 0.034 | 0.0406 | 0.402 | [-0.046, 0.114] |
| **PEQ Victimization x Gender diversity (Most GD)** | **0.064** | **0.0365** | **0.081** | **[-0.008, 0.135]** |

**Supplementary Table 12. In the most compared to least gender diverse group, there is no difference in the effect of bullying victimization (PEQ-Victimization) on broad mental health (BPM total; see PEQ Victimization x Felt gender (Most GD) term).** Statistics from linear mixed effects model regressing BPM total (broad mental health problems; dependent variable) on PEQ-Vic (bullying), gender diversity groups, and the interaction between the two. Effects of primary interest in bold. Fixed-effects covariates were age, sex, puberty, education, and family income. Random intercepts were allowed by family within site. Referent groups: gender diversity (Least GD); birth-assigned sex (female). Units: PEQ Victimization (scaled; mean = 0, sd = 1); age (years); puberty (scaled; mean = 0, standard deviation = 1); parental education (number of education years completed); family income, 10 levels ranging from 1 (< $5,000 yearly) to 10 (> $200,000 yearly).

**Supplementary Table 13.**

| **Dependent Variables:** | **PQ-BC** | | | **PEQ-Vic** | | |
| --- | --- | --- | --- | --- | --- | --- |
|  | ß | p | 95% CI | ß | p | 95% CI |
| ***State policy (high vs. low)***  *Entire sample* | -0.02 | 0.60 | [-0.10, 0.06] | -0.11 | 0.07 | [-0.22, 0.002] |
| ***State policy (high vs. low) x gender diversity (Most vs. Least)***  *Entire sample* | -0.05 | 0.59 | [-0.24,0.14] | -0.02 | 0.86 | [-0.24, 0.20] |
| ***State policy (high vs. low)***  *Most GD-only* | -0.07 | 0.73 | [-0.49, 0.34] | -0.12 | 0.54 | [-0.51, 0.26] |

**Supplementary Table 13. Null effects of cross-sectional state policy on psychotic-like experiences (PQ-BC) and bullying victimization (PEQ-Vic) at a single timepoint.** Primary interpreted effects from linear mixed effects models with PQ-BC (left) and PEQ-Vic (right) as dependent variables. Reported statistics represent the effect size, p-value, and confidence interval associated with the independent variable of interest in each model. First row: effect of state policy (ref: low support) on dependent variable in the entire sample. Second row: interaction effect between state policy (ref: low support) and gender diversity (ref: Least GD) on dependent variable in the entire sample. Third row: effect of state policy on dependent variable in the most gender diverse group only. Thus, statistics from 6 models are represented here. Fixed-effects covariates include age, birth-assigned sex, pubertal status, parental education, family income, and state-level Gini coefficient. A random intercept is allowed for family within site.

**Supplementary Table 14.**

| **Term (DV = PQ-BC)** | **Estimate** | **Std.Error** | **P** | **95% CI** |
| --- | --- | --- | --- | --- |
| (Intercept) | 0.55 | 0.5611 | 0.332 | [-0.549, 1.65] |
| Gender diversity (1-STEP) | 0.21 | 0.0635 | 9.4e-04 | [0.086, 0.335] |
| Gender diversity (2-STEP) | 0.379 | 0.079 | 1.67e-06 | [0.224, 0.534] |
| Gender diversity (Most GD) | 0.459 | 0.069 | 3.36e-11 | [0.323, 0.594] |
| State policy (High support) | 0.02 | 0.0405 | 0.621 | [-0.059, 0.1] |
| PEQ Victimization | 0.199 | 0.0215 | 2.72e-20 | [0.157, 0.242] |
| Age | -0.054 | 0.0212 | 0.011 | [-0.096, -0.013] |
| Birth-assigned sex (Male) | -0.012 | 0.0314 | 0.708 | [-0.073, 0.05] |
| Puberty | 0.05 | 0.0164 | 0.002 | [0.018, 0.082] |
| Parental education | -0.024 | 0.007 | 6.3e-04 | [-0.038, -0.01] |
| Family income | -0.023 | 0.0075 | 0.002 | [-0.037, -0.008] |
| Gini Coefficient | 1.28 | 1.0037 | 0.215 | [-0.688, 3.247] |
| Gender diversity (1-STEP) x State policy (High support) | -0.044 | 0.0897 | 0.62 | [-0.22, 0.131] |
| Gender diversity (2-STEP) x State policy (High support) | 0 | 0.1183 | 0.998 | [-0.232, 0.232] |
| Gender diversity (Most GD) x State policy (High support) | 0.059 | 0.0958 | 0.538 | [-0.129, 0.247] |
| Gender diversity (1-STEP) x PEQ Victimization | 0.118 | 0.0644 | 0.068 | [-0.008, 0.244] |
| Gender diversity (2-STEP) x PEQ Victimization | 0.04 | 0.0642 | 0.537 | [-0.086, 0.166] |
| Gender diversity (Most GD) x PEQ Victimization | 0.198 | 0.0509 | 1.02e-04 | [0.098, 0.298] |
| State policy (High support) x PEQ Victimization | 0.001 | 0.0315 | 0.965 | [-0.06, 0.063] |
| Gender diversity (1-STEP) x State policy (High support) x PEQ Victimization | 0.092 | 0.1005 | 0.358 | [-0.105, 0.289] |
| Gender diversity (2-STEP) x State policy (High support) x PEQ Victimization | 0.153 | 0.0986 | 0.12 | [-0.04, 0.346] |
| **Gender diversity (Most GD) x State policy (High support) x PEQ Victimization** | **-0.27** | **0.0764** | **4.11e-04** | **[-0.42, -0.12]** |

**Supplementary Table 14. The effect of bullying victimization (PEQ-Victimization) on psychotic-like experiences (PQ-BC) is weaker in the most gender diverse participants in states with supportive policies.** Statistics from linear mixed effects model regressing PQ-BC (psychotic-like experiences; dependent variable) on gender diversity group, state policy, bullying victimization and the interaction between the three. Effects of primary interest in bold. Fixed-effects covariates included were age, sex, puberty, parental education, combined family income, and the state-level Gini coefficient. A random intercept for family within site are modeled. Referent groups: gender diversity (Least GD); state policy (low support); birth-assigned sex (female). Units: PEQ Victimization (scaled; mean = 0, sd = 1); age (years); puberty (scaled; mean = 0, standard deviation = 1); parental education (number of education years completed); family income, 10 levels ranging from 1 (< $5,000 yearly) to 10 (> $200,000 yearly); Gini coefficient, continuous variable ranging from 0 (perfect income equality) to 1 (perfect income inequality).

**Supplementary Table 15.**

| **Term (DV = PQ-BC)** | **Estimate** | **Std.Error** | **P** | **95% CI** |
| --- | --- | --- | --- | --- |
| (Intercept) | 0.993 | 2.9714 | 0.74 | [-4.831, 6.817] |
| State policy (High support) | 0.052 | 0.1986 | 0.797 | [-0.338, 0.441] |
| PEQ Victimization | 0.363 | 0.0821 | 1.4e-05 | [0.202, 0.524] |
| Age | -0.084 | 0.1262 | 0.508 | [-0.331, 0.164] |
| Birth-assigned sex (Male) | -0.056 | 0.2458 | 0.819 | [-0.538, 0.425] |
| Puberty | 0.165 | 0.1167 | 0.159 | [-0.064, 0.394] |
| Parental education | -0.035 | 0.0437 | 0.421 | [-0.121, 0.05] |
| Family income | -0.02 | 0.0417 | 0.639 | [-0.101, 0.062] |
| Gini Coefficient | 2.365 | 5.2153 | 0.655 | [-7.856, 12.587] |
| **State policy (High support) x PEQ Victimization** | **-0.202** | **0.1245** | **0.106** | **[-0.446, 0.042]** |

**Supplementary Table 15. The effect of bullying (PEQ Victimization) on psychotic-like experiences (PQ-BC) is no different in the most gender diverse participants in low-support states compared to the most gender diverse participants in high-support states.** Statistics from linear mixed effects model regressing PQ-BC (psychotic-like experiences; dependent variable) on state policy, bullying victimization and the interaction between the two in the most gender diverse group only (N=689). Fixed-effects covariates included were age, birth-assigned sex, puberty, parental education, combined family income, and the state-level Gini coefficient. A random intercept for family within site as modeled. Effects of primary interest in bold. Referent groups: state policy (low support); birth-assigned sex (female). Units: PEQ Victimization (scaled; mean = 0, sd = 1); age (years); puberty (scaled; mean = 0, standard deviation = 1); parental education (number of education years completed); family income, 10 levels ranging from 1 (< $5,000 yearly) to 10 (> $200,000 yearly); Gini coefficient, continuous variable ranging from 0 (perfect income equality) to 1 (perfect income inequality). *This table differs from the* ***Supplementary Table 14*** *in that the referent group for the interaction term is the least gender diverse group in low support states.*

**Supplementary Table 16.**

| **Term (DV = PQ-BC)** | **Estimate** | **Std.Error** | **P** | **95% CI** |
| --- | --- | --- | --- | --- |
| (Intercept) | 0.906 | 0.0699 | 5.47e-26 | [0.769, 1.043] |
| **Time** | **-0.103** | **0.0129** | **2.18e-08** | **[-0.129, -0.078]** |
| Birth-assigned sex (Male) | -0.084 | 0.0157 | 9.97e-08 | [-0.114, -0.053] |
| Puberty | 0.048 | 0.0077 | 3.36e-10 | [0.033, 0.063] |
| Parental education | -0.022 | 0.004 | 6.77e-08 | [-0.029, -0.014] |
| Family income | -0.036 | 0.0043 | 1.36e-16 | [-0.044, -0.027] |

**Supplementary Table 16. On average, across the entire sample, rates of psychotic-like experiences (PQ-BC) declined over time.** Statistics from longitudinal, linear mixed effects model regressing PQ-BC (psychotic-like experiences; dependent variable) on time point. Fixed-effects covariates included were birth-assigned sex, puberty, parental education, and combined family income. The model included a random intercept for subject within family within site and allowed the effect of time to vary (random slope). Effects of primary interest in bold. Referent groups: birth-assigned sex (female). Units: time (data collection wave, 1-4); puberty (scaled; mean = 0, standard deviation = 1); parental education (number of education years completed); family income, 10 levels ranging from 1 (< $5,000 yearly) to 10 (> $200,000 yearly).

**Supplementary Table 17.**

| **Term (DV = PQ-BC)** | **Estimate** | **Std.Error** | **P** | **95% CI** |
| --- | --- | --- | --- | --- |
| (Intercept) | 0.764 | 0.0681 | 3.61e-21 | [0.63, 0.897] |
| Gender diversity (1-STEP) | 0.225 | 0.0421 | 9.03e-08 | [0.142, 0.307] |
| Gender diversity (2-STEP) | 0.304 | 0.0592 | 2.88e-07 | [0.188, 0.42] |
| Gender diversity (Most GD) | 0.473 | 0.0529 | 4.52e-19 | [0.369, 0.576] |
| Time | -0.107 | 0.0127 | 5.25e-09 | [-0.132, -0.082] |
| Birth-assigned sex (Male) | -0.003 | 0.0154 | 0.843 | [-0.033, 0.027] |
| Puberty | 0.04 | 0.0075 | 8.69e-08 | [0.026, 0.055] |
| Parental education | -0.021 | 0.0038 | 3.52e-08 | [-0.029, -0.014] |
| Family income | -0.032 | 0.0041 | 2.4e-14 | [-0.04, -0.024] |
| Gender diversity (1-STEP) x Time | -0.025 | 0.0167 | 0.139 | [-0.058, 0.008] |
| Gender diversity (2-STEP) x Time | 0.011 | 0.0227 | 0.622 | [-0.033, 0.056] |
| **Gender diversity (Most GD) x Time** | **0.059** | **0.0189** | **0.002** | **[0.022, 0.096]** |

**Supplementary Table 17. Regardless of state policies at a single time point, PLEs declined more slowly in the most gender diverse participants (Most GD).** Statistics from linear mixed effects model regressing PQ-BC (psychotic-like experiences; dependent variable) on gender diversity group, time point, and the interaction between the two. Fixed-effects covariates included were birth-assigned sex, puberty, parental education, and combined family income. The model included a random intercept for subject within family within site and allowed the effect of time to vary (random slope). Effects of primary interest in bold. Referent groups: gender diversity (Least GD); birth-assigned sex (female). Units: time (data collection wave, 1-4); puberty (scaled; mean = 0, standard deviation = 1); parental education (number of education years completed); family income, 10 levels ranging from 1 (< $5,000 yearly) to 10 (> $200,000 yearly); Gini coefficient, continuous variable ranging from 0 (perfect income equality) to 1 (perfect income inequality).

**Supplementary Table 18.**

| **Term (DV = PQ-BC)** | **Estimate** | **Std.Error** | **P** | **95% CI** |
| --- | --- | --- | --- | --- |
| (Intercept) | -0.074 | 0.5274 | 0.889 | [-1.107, 0.96] |
| Gender diversity (1-STEP) | 0.309 | 0.0937 | 9.67e-04 | [0.126, 0.493] |
| Gender diversity (2-STEP) | 0.254 | 0.1268 | 0.045 | [0.005, 0.503] |
| Gender diversity (Most GD) | 0.983 | 0.1316 | 8.63e-14 | [0.726, 1.241] |
| Time | -0.057 | 0.0193 | 0.006 | [-0.095, -0.019] |
| State policy (Increasing) | 0.145 | 0.1056 | 0.178 | [-0.062, 0.352] |
| State policy (Consistently low) | 0.11 | 0.1072 | 0.316 | [-0.1, 0.32] |
| Birth-assigned sex (Male) | -0.036 | 0.0229 | 0.115 | [-0.081, 0.009] |
| Puberty | 0.027 | 0.0121 | 0.025 | [0.003, 0.051] |
| Parental education | -0.019 | 0.0057 | 0.001 | [-0.03, -0.007] |
| Family income | -0.03 | 0.0062 | 9.13e-07 | [-0.043, -0.018] |
| Gini Coefficient | 1.364 | 1.082 | 0.215 | [-0.757, 3.485] |
| Gender diversity (1-STEP) x Time | -0.046 | 0.0336 | 0.167 | [-0.112, 0.019] |
| Gender diversity (2-STEP) x Time | 0.015 | 0.0446 | 0.73 | [-0.072, 0.103] |
| Gender diversity (Most GD) x Time | -0.083 | 0.0393 | 0.036 | [-0.16, -0.006] |
| Gender diversity (1-STEP) x State policy (Increasing) | -0.067 | 0.148 | 0.652 | [-0.357, 0.223] |
| Gender diversity (2-STEP) x State policy (Increasing) | -0.287 | 0.2011 | 0.154 | [-0.681, 0.107] |
| Gender diversity (Most GD) x State policy (Increasing) | -0.307 | 0.223 | 0.169 | [-0.744, 0.13] |
| Gender diversity (1-STEP) x State policy (Consistently low) | -0.077 | 0.1395 | 0.58 | [-0.351, 0.196] |
| Gender diversity (2-STEP) x State policy (Consistently low) | 0.144 | 0.1993 | 0.471 | [-0.247, 0.534] |
| Gender diversity (Most GD) x State policy (Consistently low) | -0.932 | 0.1815 | 2.84e-07 | [-1.288, -0.577] |
| Time x State policy (Increasing) | -0.046 | 0.0276 | 0.102 | [-0.1, 0.008] |
| Time x State policy (Consistently low) | -0.036 | 0.0271 | 0.191 | [-0.09, 0.017] |
| Gender diversity (1-STEP) x Time x State policy (Increasing) | -0.013 | 0.0533 | 0.802 | [-0.118, 0.091] |
| Gender diversity (2-STEP) x Time x State policy (Increasing) | 0.113 | 0.0703 | 0.107 | [-0.024, 0.251] |
| Gender diversity (Most GD) x Time x State policy (Increasing) | 0.076 | 0.0669 | 0.254 | [-0.055, 0.207] |
| Gender diversity (1-STEP) x Time x State policy (Consistently low) | 0.019 | 0.0515 | 0.714 | [-0.082, 0.12] |
| Gender diversity (2-STEP) x Time x State policy (Consistently low) | -0.056 | 0.0696 | 0.423 | [-0.192, 0.081] |
| **Gender diversity (Most GD) x Time x State policy (Consistently low)** | **0.317** | **0.0566** | **2.15e-08** | **[0.206, 0.428]** |

**Supplementary Table 18. Psychotic-like experiences differentially increased over time in the most gender diverse participants in consistently low support states.** Statistics from linear mixed effects model regressing PQ-BC (psychotic-like experiences; dependent variable) on gender diversity group, time point, longitudinal state-level policy, and their interactions. Effects of primary interest in bold. Fixed-effects covariates included were birth-assigned sex, puberty, parental education, combined family income, and Gini coefficient. The model included a random intercept for subject within family within site and allowed the effect of time to vary (random slope). Referent groups: gender diversity (Least GD); birth-assigned sex (female); state policy (Consistently High). Units: time (data collection wave, 1-4); puberty (scaled; mean = 0, standard deviation = 1); parental education (number of education years completed); family income, 10 levels ranging from 1 (< $5,000 yearly) to 10 (> $200,000 yearly); Gini coefficient, continuous variable ranging from 0 (perfect income equality) to 1 (perfect income inequality).

SUPPLEMENTARY TABLES CONTINUED ON NEXT PAGE

**Supplementary Table 19.**

| **Term (DV = PQ-BC)** | **Estimate** | **Std.Error** | **P** | **95% CI** |
| --- | --- | --- | --- | --- |
| (Intercept) | 1.375 | 2.3131 | 0.558 | [-3.158, 5.909] |
| Time | -0.188 | 0.0875 | 0.032 | [-0.359, -0.016] |
| State policy (Increasing) | 0.022 | 0.4161 | 0.958 | [-0.793, 0.837] |
| State policy (Consistently low) | -0.907 | 0.3418 | 0.009 | [-1.577, -0.237] |
| Birth-assigned sex (Male) | -0.163 | 0.1962 | 0.405 | [-0.548, 0.221] |
| Puberty | 0.069 | 0.0944 | 0.465 | [-0.116, 0.254] |
| Parental education | -0.049 | 0.0385 | 0.202 | [-0.125, 0.026] |
| Family income | -0.026 | 0.0388 | 0.502 | [-0.102, 0.05] |
| Gini Coefficient | 1.836 | 4.5618 | 0.692 | [-7.105, 10.777] |
| Time x State policy (Increasing) | -0.007 | 0.1263 | 0.955 | [-0.255, 0.24] |
| **Time x State policy (Consistently low)** | **0.298** | **0.1078** | **0.006** | **[0.087, 0.51]** |

**Supplementary Table 19. In the Most GD group in states with consistently unsupportive policies related to gender identity, the change in PQ-BC over time was greater than in the Most GD participants in states with consistently supportive policies.** Statistics from linear mixed effects model regressing PQ-BC (psychotic-like experiences; dependent variable) on time point, state policy, and the interaction between the two in the sample of most gender diverse (Most GD) participants. Fixed-effects covariates included were birth-assigned sex, puberty, parental education, and combined family income. The model included a random intercept for subject within family within site and allowed the effect of time to vary (random slope). Effects of primary interest in bold. Referent groups: state policy (Consistently high); birth-assigned sex (female). Units: time (data collection wave, 1-4); puberty (scaled; mean = 0, standard deviation = 1); parental education (number of education years completed); family income, 10 levels ranging from 1 (< $5,000 yearly) to 10 (> $200,000 yearly); Gini coefficient, continuous variable ranging from 0 (perfect income equality) to 1 (perfect income inequality).

**Supplementary Table 20**

|  | **ß** | **95% CI** | **P** |
| --- | --- | --- | --- |
| **High support, no change (H-H)** | | | |
| Least Gender Diverse | -0.010 | [-0.021, 0.001] | 0.086 |
| 1-step | -0.005 | [-0.108, 0.097] | 0.919 |
| 2-step | 0.013 | [-0.103, 0.129] | 0.827 |
| Most Gender Diverse | -0.060 | [-0.198, 0.077] | 0.387 |
| **Increased support** | | | |
| Least Gender Diverse | -0.014 | [-0.026, -0.002] | 0.019 |
| 1-step | -0.040 | [-0.092, 0.011] | 0.123 |
| 2-step | 0.111 | [-0.041, 0.262] | 0.149 |
| Most Gender Diverse | -0.074 | [-0.231, 0.082] | 0.350 |
| **Low support, no change (L-L)** | | | |
| Least Gender Diverse | -0.016 | [-0.028, -0.004] | 0.011 |
| 1-step | -0.050 | [-0.128, 0.027] | 0.203 |
| 2-step | -0.005 | [-0.164, 0.154] | 0.954 |
| Most Gender Diverse | 0.184 | [0.062, 0.306] | 3.42e-04* |
| * represents significance after FDR correction for multiple comparisons | | | |

**Supplementary Table 20. Psychotic-like experiences show significant changes over time only.** Effect of time point on PQ-BC score in each gender diversity group stratified by state policy over time. Each row represents statistics from a model conducted in a gender diversity group (e.g., the last row represents the effect of time on PQ-BC in the most gender diverse participants in consistently unsupportive states. Models are adjusted for birth-assigned sex, pubertal status (time-variant), parental education, and family income and preserve the nested structure of the main interaction model (i.e., random slopes by time nested within subject, within family and within study site). A positive coefficient represents a positive change (increase) in PQ-BC scores over time. Asterisks indicate significance after correction for multiple comparisons.

**Supplementary Table 21.**

***Negative Laws***

| **Level of State Law** | **Corresponding Point Value** |
| --- | --- |
| State bans transgender people from using bathrooms and facilities consistent with their gender identity in K-12 schools and at least some government-owned buildings (e.g., public colleges/universities, prisons or jails, etc). | -0.5 |
| State bans transgender people from using bathrooms and facilities consistent with their gender identity in all government-owned buildings and spaces, including K-12 schools, colleges, and more. | -1 |

***Positive Laws***

| **Level of State Law** | **Corresponding Point Value** |
| --- | --- |
| No state law prohibiting employment discrimination based on gender identity, but 50-99% of state population is protected from discrimination based on gender identity through local ordinances. | +0.5 |
| State law explicitly prohibits employment discrimination based on gender identity. | +1 |

**Supplementary Table 21.** From the Movement Advancement Project website providing an example of two laws that would receive a value of -1 vs -0.5 (top; unsupportive laws) and 1 vs 0.5 (bottom; supportive laws).
